# Gaussian Process Emulation for Exploring Complex Infectious Disease Models

**DOI:** 10.1101/2024.11.28.24318136

**Authors:** Anna M. Langmüller, Kiran A. Chandrasekher, Benjamin C. Haller, Samuel E. Champer, Courtney C. Murdock, Philipp W. Messer

**Affiliations:** Department of Computational Biology, Cornell University, Ithaca, NY 14853, USA; Department of Mathematics, University of Vienna, Vienna 1090, Austria; Aarhus Institute of Advanced Studies, Aarhus University, Aarhus C 8000, Denmark; Department of Entomology, Cornell University, Ithaca, NY 14853, USA; Cornell Institute of Host-Microbe Interactions and Disease, Cornell University, Ithaca, NY 14853, USA; Center for the Ecology of Infectious Diseases, University of Georgia, Athens GA, USA

**Keywords:** individual-based modeling, statistical emulation, Gaussian Processes, epidemiological modeling, variance-based sensitivity analysis

## Abstract

Epidemiological models that aim for a high degree of biological realism by simulating every individual in a population are unavoidably complex, with many free parameters, which makes systematic explorations of their dynamics computationally challenging. This study investigates the potential of Gaussian Process emulation to overcome this obstacle. To simulate disease dynamics, we developed an abstract individual-based model that is loosely inspired by dengue, incorporating some key features shaping dengue epidemics such as social structure, human movement, and seasonality. We trained three Gaussian Process surrogate models on three outcomes: outbreak probability, maximum incidence, and epidemic duration. These surrogate models enable the rapid prediction of outcomes at any point in the eight-dimensional parameter space of the original model. Our analysis revealed that average infectivity and average human mobility are key drivers of these epidemiological metrics, while the seasonal timing of the first infection can influence the course of the epidemic outbreak. We use a dataset comprising more than 1,000 dengue epidemics observed over 12 years in Colombia to calibrate our Gaussian Process model and evaluate its predictive power. The calibrated Gaussian Process model identifies a subset of municipalities with consistently higher average infectivity estimates, which show notable overlap with previously reported dengue disease clusters, suggesting that statistical emulation can facilitate empirical data analysis. Overall, this work underscores the potential of Gaussian Process emulation to enable the use of more complex individual-based models in epidemiology, allowing a higher degree of realism and accuracy that should increase our ability to control diseases of public health concern.

**Author Summary:** Detailed individual-based models can capture a high degree of realism, but their complexity often makes them too slow or cumbersome to explore fully. In our work, we explore how Gaussian Process emulation — a statistical method for building fast, accurate surrogate models — can help overcome this challenge. First, we developed an individual-based model that simulates disease spread in a population, accounting for features such as social structure, human mobility, and seasonal variation in infection risk. We then trained a Gaussian Process surrogate model on the outputs of this individual-based model, which allowed us to predict key outcomes almost instantly across a wide range of parameter values. This approach made it possible to systematically explore which factors drive simulated epidemics. We found that two variables — average infectivity and average mobility — had the greatest influence on whether and how outbreaks occurred. Our results demonstrate that Gaussian Process emulation offers a practical and powerful way to study complex disease systems. While we applied this approach to infectious disease transmission, the underlying method can be useful for analyzing many other types of detailed, simulation-based models.

## Introduction

Simulation models that describe individual organisms or agents have become a well-established research tool across numerous scientific fields [1]. These so-called individual-based models (IBMs) allow researchers to explore how system-level characteristics emerge from individual behaviors, while also investigating the reciprocal influence of the system on individuals [1]. In the field of epidemiology, IBMs have provided valuable insights into the dynamics of pathogen and disease spread and have facilitated rigorous evaluation of planned intervention strategies, making them an integral part of modern epidemiological research [2–6]. Recent computational advances, combined with the development of comprehensive individual-based simulation frameworks [2,7], have enabled the creation of epidemiological models with unprecedented realism. Such models can capture a range of details, including, for example, fine-scale human movement [5,8] and specific larval breeding sites for organisms relevant to vector-borne disease transmission [3].

While enhanced biological realism in IBMs has undeniably deepened our understanding of epidemiological processes, it also introduces increased complexity because of the level of detail being simulated, which comes at a substantial computational cost. As IBMs become more realistic, they also become more parameter-rich, making it increasingly difficult to identify the key drivers of disease dynamics. This is partly because parameters often interact in complex, non-linear ways, complicating efforts to quantify the contribution of any single factor to model outcomes. Global sensitivity analysis can help by quantifying the relative contribution of each parameter — as well as their interactions — to IBM outcomes. For example, the Sobol method [9] is a global sensitivity analysis approach that is able to assess complex, non-linear parameter interactions by partitioning the observed variance in IBM output into relative contributions from single parameters as well as interactions between two or more parameters. This allows researchers to gain a deeper understanding of the key drivers of the disease dynamics observed in a simulation.

Global sensitivity analysis can provide valuable insights into model dynamics, but generating sufficient data for robust sensitivity analysis can be computationally demanding, especially for IBMs. The computational demand of IBMs typically scales at least linearly with population size and can increase quadratically when intricate behaviors or pairwise interactions are modeled. Unlike mechanistic models based on ordinary differential equations [10], which can often be analyzed analytically or with relatively modest computational effort, IBMs often require thousands of simulation runs to fully explore their complex parameter spaces. This makes comprehensive parameter space exploration particularly challenging for high-dimensional IBMs, even when they are optimized for runtime performance, quickly leading to the well-known “Curse of Dimensionality” [11].

Statistical emulation [11] can address this problem by constructing a fast, predictive surrogate model based on a limited set of IBM simulations. Once trained, the emulator can predict the IBM’s outputs across the full parameter space in a fraction of the time it would take to generate the same outputs with the IBM. This allows efficient execution of tasks such as sensitivity analysis, model output exploration, and model calibration, at a resolution that would be infeasible using the IBM directly. In essence, statistical emulation facilitates the extraction of meaningful insights from complex IBMs by substantially reducing the computational costs associated with their analysis. A well-designed emulator can reduce computational runtimes from days to mere seconds, dramatically expanding the scope of epidemiological modeling. While emulators often rely on statistical methods, recent advances increasingly incorporate machine learning techniques [12].

Gaussian Processes (GPs), first introduced in the 1960s within the field of geostatistics [13,14], are flexible statistical emulators that have been successfully applied across a range of disciplines [15–17]. GPs are non-parametric models that define a distribution over functions based on observed data. A key advantage of GPs over other machine learning techniques, such as conventional support vector machines or neural networks, lies in their Bayesian foundations, which allow GPs to provide confidence intervals alongside their predictions. This uncertainty quantification enables efficient sampling of additional training data from regions with the greatest uncertainty, facilitating active learning [18] that can quickly produce highly accurate emulation. Furthermore, with the availability of advanced software packages supporting GPU acceleration, the computational efficiency of GPs has improved at an astonishing pace in recent years, making them an increasingly attractive research tool [19].

In epidemiology, the ability of GPs to efficiently extrapolate between sparse data points is often utilized for estimating disease incidence counts in areas where data is missing or unobserved [20,21]. GPs also serve as valuable forecasting tools [17,22], and are key components of early-warning systems [23]. Furthermore, as emulators of complex, computationally intensive IBMs, GPs facilitate the calibration of these IBMs to empirical data by helping to select parameter values for which the IBM’s outputs closely match observed real-world data [24–26]. Notable examples of using GPs as emulators to better understand complex IBMs include recent studies that applied GP emulation to the OpenMalaria model (an advanced IBM developed to simulate malaria transmission and control) [26] to explore key drivers of the spread of drug-resistant *Plasmodium falciparum* [27], and to assess the effectiveness of various malaria intervention strategies [28,29]. While these applications demonstrate the power of GP emulation, they are often closely tied to specific contexts and require a deep familiarity with the particular disease system.

In this study, we developed an abstract, generalizable IBM to highlight the potential of GP emulation in epidemiological modeling. Specifically, we focus on variance-based sensitivity analysis across a high-dimensional parameter space to illustrate how GP emulation can be used not only to accelerate computations, but also to improve our understanding of complex epidemiological models. First, we developed an abstract individual-based disease transmission model that is loosely inspired by dengue, incorporating some key features shaping dengue epidemics such as social structure, human movement, and seasonality. Dengue poses a growing global health threat [30], with cases rapidly increasing due to urbanization [31] and climate change that has expanded the habitat of *Aedes* mosquitoes, the primary vectors of the dengue virus [32,33]. Next, we employed GP surrogate models to efficiently predict three key metrics from the IBM: outbreak probability, maximum incidence, and duration of outbreaks.

We demonstrate that our GPs explore the parameter space with impressive speed, enabling comprehensive sensitivity analysis. We conducted a global sensitivity analysis and found that average infectivity and average human mobility are primary drivers of outbreak dynamics, while the interactions between seasonality strength and initial infection timing can critically influence the course of epidemic outbreaks. To determine whether insights from our sensitivity analysis could deepen our understanding of real-world epidemics, we investigated weekly dengue incidence data at the municipality level across Colombia over more than a decade [34,35], uncovering a subset of municipalities with consistently high infectivity estimates that showed notable overlap with previously reported dengue disease clusters [36]. This study illustrates how statistical emulation can support complex model analysis and potentially advance their application in epidemiology.

## Results

### Individual-based model

We implemented an IBM in C++ that simulates and tracks disease transmission to study how disease dynamics are influenced by infection probability, human movement, and social structure: three key features that shape dengue epidemics [37–39]. The model is designed not to replicate a specific empirical system, but to illustrate the relative importance of these parameters and their interactions in influencing the course of epidemic outbreaks. A detailed description of the IBM can be found in the Materials & Methods section. All model parameters are summarized in Table 1. Briefly, each simulation begins by generating 10,000 locations that are each home to a group of susceptible individuals. The number of individuals per location is sampled from a negative binomial distribution fitted to the demography of Iquitos, Peru — a well-studied dengue transmission hotspot [37,40]. These locations are then randomly organized into non-overlapping family clusters, with the “family cluster size” parameter controlling the number of locations per cluster. Social structure, controlled by the “social structure” parameter, influences the likelihood that individuals interact within their family cluster. Human movement — the number of visits to locations per day — is sampled from a negative binomial distribution, defined by the “average mobility” and “mobility skewness” parameters. The social structure of the model is depicted in Supplemental Figure S1.

**Table 1.** Parameters of the individual-based disease transmission model.

| Parameter | Description | Default | Range |
| --- | --- | --- | --- |
| Average infectivity | The average infection probability across a year, which removes the effect of seasonality | 0.015 | [0, 0.03] |
| Seasonality strength | A scaling factor between 0 and 1 that controls the magnitude of seasonal variation in infection probability | 0.5 | [0, 1] |
| First case timing | Determines the timing of the first case relative to the seasonal peak in infection probability | 0 | [0, 1] |
| Infectious period | The average number of days an individual remains infectious, with actual days determined by probabilistically rounding to the nearest integers around the specified value | 5 | [4, 6] |
| Average mobility | The average number of visits a human makes to locations per day, in addition to their family home | 2 | [1, 5] |
| Mobility skewness | The success probability in the negative binomial distribution that determines the number of visits a human makes per day; a lower value results in greater variance in the daily visit count | 0.5 | [0.05, 0.95] |
| Social structure | The probability of a visit occurring within the family cluster of the individual moving | 0.5 | [0, 1] |
| Family cluster size | The average number of locations assigned to each cluster, with actual sizes determined by probabilistically rounding to the nearest integers around the specified value | 5 | [1, 20] |

The disease is introduced by infecting a single randomly chosen individual. Infected individuals remain contagious for a number of days specified by the “infectious period” parameter, after which they recover and gain lasting immunity (and thus cannot become reinfected). When a susceptible individual visits a location that was visited by infectious individuals the day before, the likelihood of infection from each previous infectious visitor is determined by the infection probability. In the context of dengue, seasonal fluctuations in this infection probability can be interpreted as reflecting changes in mosquito abundance over the year. Although dengue is transmitted by mosquitoes, we do not model individual mosquitoes in our abstract simulation framework. This choice was driven by the very limited dispersal ability of *Aedes aegypti* [41], the primary vector for dengue in the Americas, which predominantly bites during daylight hours [42]. Consequently, human movement patterns tend to be more influential than mosquito movement in shaping dengue dynamics [37,39,43].

The infection probability is defined by a cosine function with three parameters: (i) the “average infectivity” parameter (α_0_), representing the average infection probability over the course of a year (365 days); (ii) the “seasonality strength” parameter (α*_season_*), controlling the magnitude of seasonal variation in infection probability; and (iii) the “first case timing” parameter (*t_first_*), defining the horizontal shift of the cosine function and thus the timing of the first case relative to the peak infection probability due to seasonality. Together, these parameters define the infection probability at any given day *t* in the year:

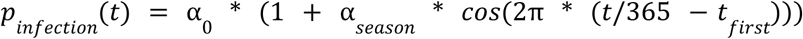

The IBM progresses by daily timesteps and continues until there are no infectious individuals left. The output consists of daily counts of individuals in each infection state (susceptible, exposed, infectious, and recovered). For each combination of parameters, we use 100 replicate simulation runs to calculate three metrics of the simulated epidemics: (i) outbreak probability, defined as the proportion of simulation runs in which more than 0.1% of the population becomes infected; (ii) maximum disease incidence (*i_max_*), defined as the highest proportion of infectious individuals seen in any timestep; and (iii) outbreak duration, defined as the timespan in days from the first infectious case to the recovery of the last infectious individual. To calculate *i_max_* and outbreak duration, we average across 100 simulation runs where an outbreak occurred (conducting additional simulations as needed to obtain 100 such runs for each parameter combination), thereby minimizing confounding effects from stochastic losses of the disease.

We systematically varied the eight parameters outlined above to explore how the simulated epidemics change across the parameter space. Across the full range of parameters (Table 1), the three metrics vary significantly: the average outbreak probability is 0.79, ranging from 0 to 1; the average *i_max_* is 0.67, ranging from 0.0003 to 0.99; and the average duration is 63.88 days, ranging from 19.65 to 424.15 days.

### Gaussian Process training & performance

Even for relatively simple IBMs, generating the necessary data for a comprehensive sensitivity analysis can become computationally prohibitive, especially when the parameter space is high-dimensional and parameters interact with one another in complex ways. To address this, we implemented GP surrogate models as statistical emulation tools [11,12], and trained them on input–output data pairs from our IBM (Materials & Methods section). We trained three independent GPs to predict outbreak probability, *i_max_*, and outbreak duration. The Bayesian nature of GPs allows for uncertainty quantification of single predictions [18], which we utilized in an active learning loop to adaptively select additional training data points (Figure 1A).

**Figure 1.**
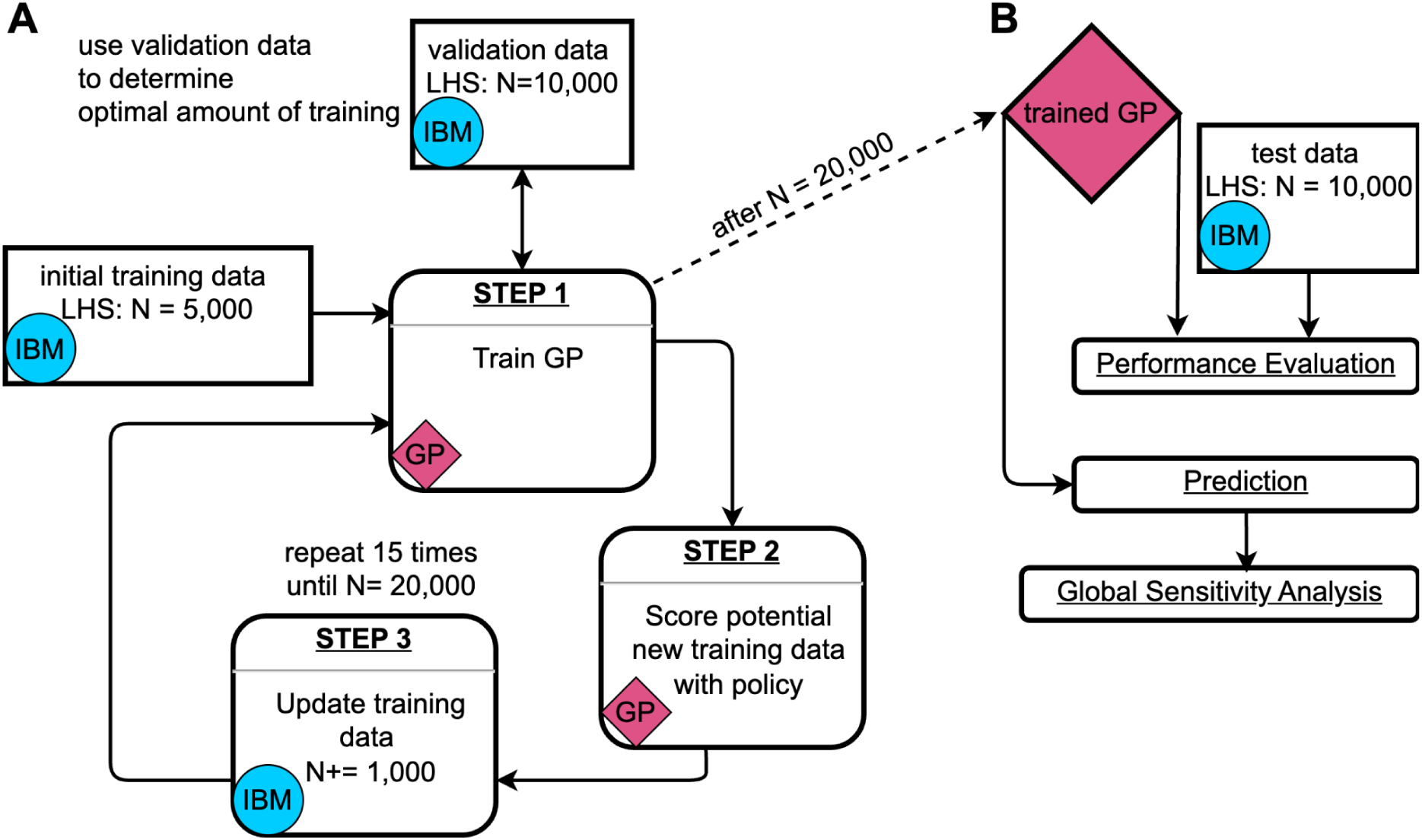
(A) Gaussian Process (GP) training loop [18]. The GP training begins with an initial training dataset consisting of a Latin hypercube sample (LHS) of 5,000 data points generated from the input domain (Table 1) using the individual-based simulation model (IBM). During training, the GP is continuously evaluated against a validation dataset of 10,000 data points to determine the optimal amount of training iterations and prevent overfitting. After each training cycle, 10^7^ potential new data points are scored based on a policy that considers their predicted value and 95% confidence interval. In each iteration of the training loop, 1,000 additional data points are sampled from those 10^7^ candidate points, where the probability of being sampled is proportional to their respective scores. The newly selected data points are then simulated using the IBM, added to the training dataset, and the next training round begins. (B) Usage of the trained GP. After training, the GP is tested using an independent dataset of 10,000 LHS data points to evaluate its performance. The trained GP can then be used for rapid predictions, enabling large-scale global sensitivity analyses.

Thanks to the optimization techniques and GPU acceleration implemented in GPyTorch [19], GP training remained computationally manageable. On our local machine (i5-12600K CPU, GeForce RTX 4090 GPU), one training round with 30,000 iterations took approximately 15 minutes to 2 hours, depending on the size of the training data (5,000 – 20,000 points, Figure 1A: Step 1). The majority of time in the overall process was spent generating additional training data with the IBM during the active learning rounds (Figure 1A: Step 3). This was more time-intensive (∼10 hours on our local machine) for the GPs modeling *i_max_*and outbreak duration than for the GP modeling outbreak probability, because the training data for the latter also included many simulation runs where no outbreaks occurred (which are typically very fast).

One of the primary goals of statistical emulation is to reduce computational time [12]. GPs are particularly efficient for this purpose because they provide closed-form solutions for predictions, and computationally expensive matrix inversions are performed during training, not during prediction [18,44]. Thus, once trained, GPs can predict new data points rapidly. In our case, we observed a significant speed-up: with our local machine we were able to predict the 10,000 data points needed for Figure 3C in only 0.1 seconds. This is equivalent to performing at least 10^6^ IBM simulation runs, which, depending on available computational resources, would typically take several hours in a computing cluster environment. Additionally, the runtime for GP predictions is deterministic and constant due to the closed-form solution, whereas the runtime of IBM simulations varies depending on the duration of the disease outbreak.

We evaluated model performance using root mean square error (RMSE) — both for continuous checks against a validation dataset during training to avoid overfitting (Figure 1A), and for assessing the accuracy of the GPs after training was completed (Figure 1B). During training, we observed that the first few adaptive training rounds tended to lead to the most significant improvements in model performance, whereas additional rounds later in the process yielded diminishing returns (Figure S2). The final GPs achieved RMSE values of 0.057 for outbreak probability, 0.042 for *i_max_*, and 0.068 for duration (Figure S2).

We observed greater variance in the model’s predictive accuracy for weaker epidemics (i.e., lower *i_max_* values, Figure 2B) and longer epidemics (i.e., larger duration, Figure 2C). In such cases, stochasticity plays a larger role, making predictions more challenging. For the *i_max_* GP, there were instances where the intensity of the epidemic outbreak was severely overpredicted (Figure 2B). This might have resulted from neighboring data points with very different properties. Although our kernel choice allows for rather abrupt changes in function values, the interpolation might not fully capture the true dynamics of the underlying model if it does not happen exactly at the midpoint between the two points.

**Figure 2.**
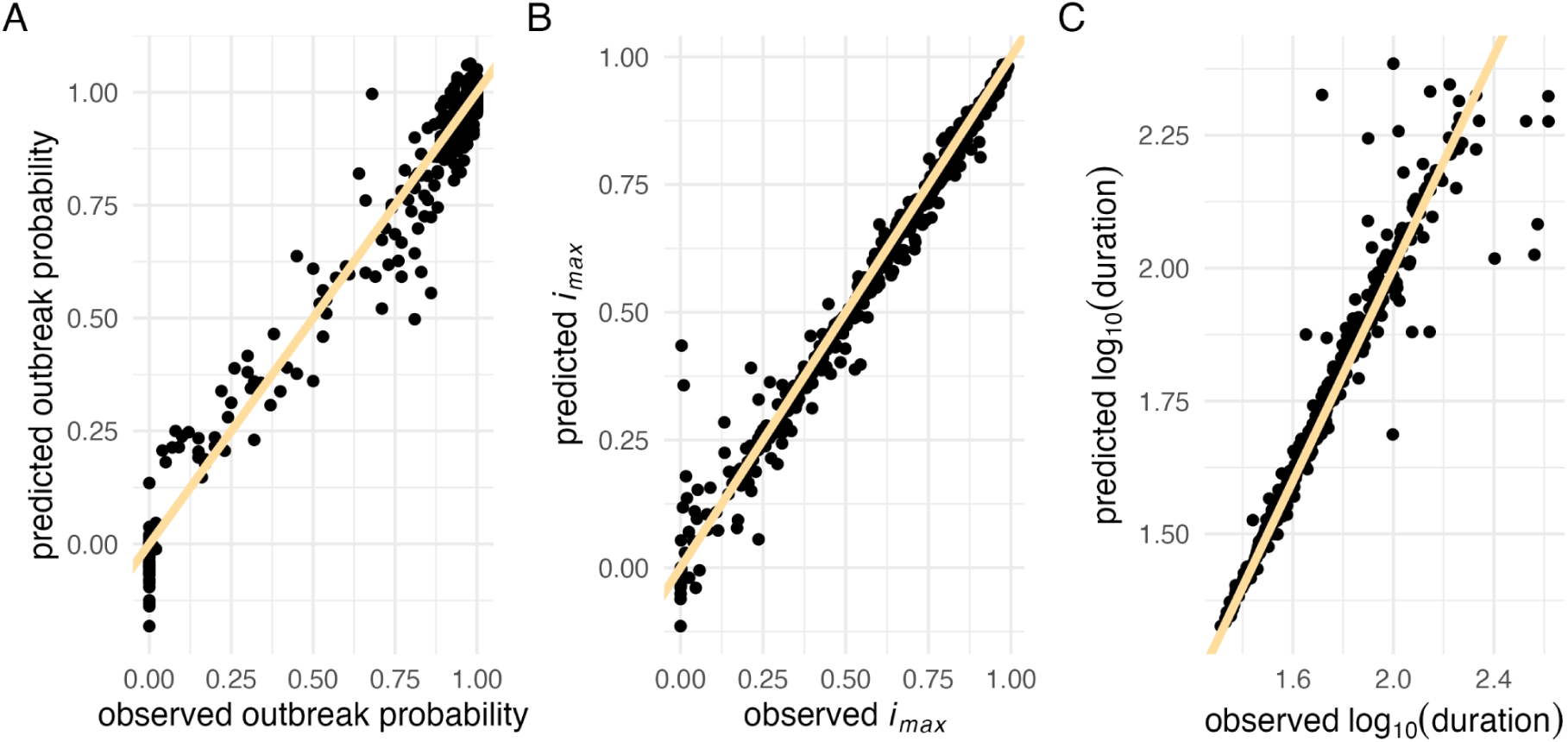
Gaussian Process performance evaluation. Comparison of observed versus predicted values for 500 randomly sampled test data points. The yellow line represents the identity line (*x = y*) for (A) outbreak probability (B) maximum incidence (*i_max_*), and (C) log_10_-transformed duration.

### Sensitivity analysis & specific model outcomes

The computational efficiency of the GPs allowed us to perform a comprehensive variance-based sensitivity analysis of our model. For this, we used the Sobol method [9], a variance-based approach that quantifies the contribution of each variable — both independently and in interaction with others — to the overall variance of the model output. We estimated first-order effects (the influence of single parameters independent of others), second-order effects (the contribution of pairwise interactions between two parameters), and total-order effects (which include first-order effects as well as all interactions of any order).

This analysis revealed that average infectivity and average mobility are the primary drivers in our model, shaping all three epidemiological metrics: outbreak probability, *i_max_*, and outbreak duration. Since there is no correlation between the number of visits sampled for a given individual over time, our model does not include systematic super-spreading behavior. As a result, we expected the sensitivity index for mobility skewness to be low across all three metrics, in contrast to a stronger impact of average mobility. Our findings confirm this expectation, with mobility skewness showing no influence on the epidemiological metrics (*i_max_*: Figure 3A, outbreak probability: Figure S3A, outbreak duration: Figure S4A).

**Figure 3.**
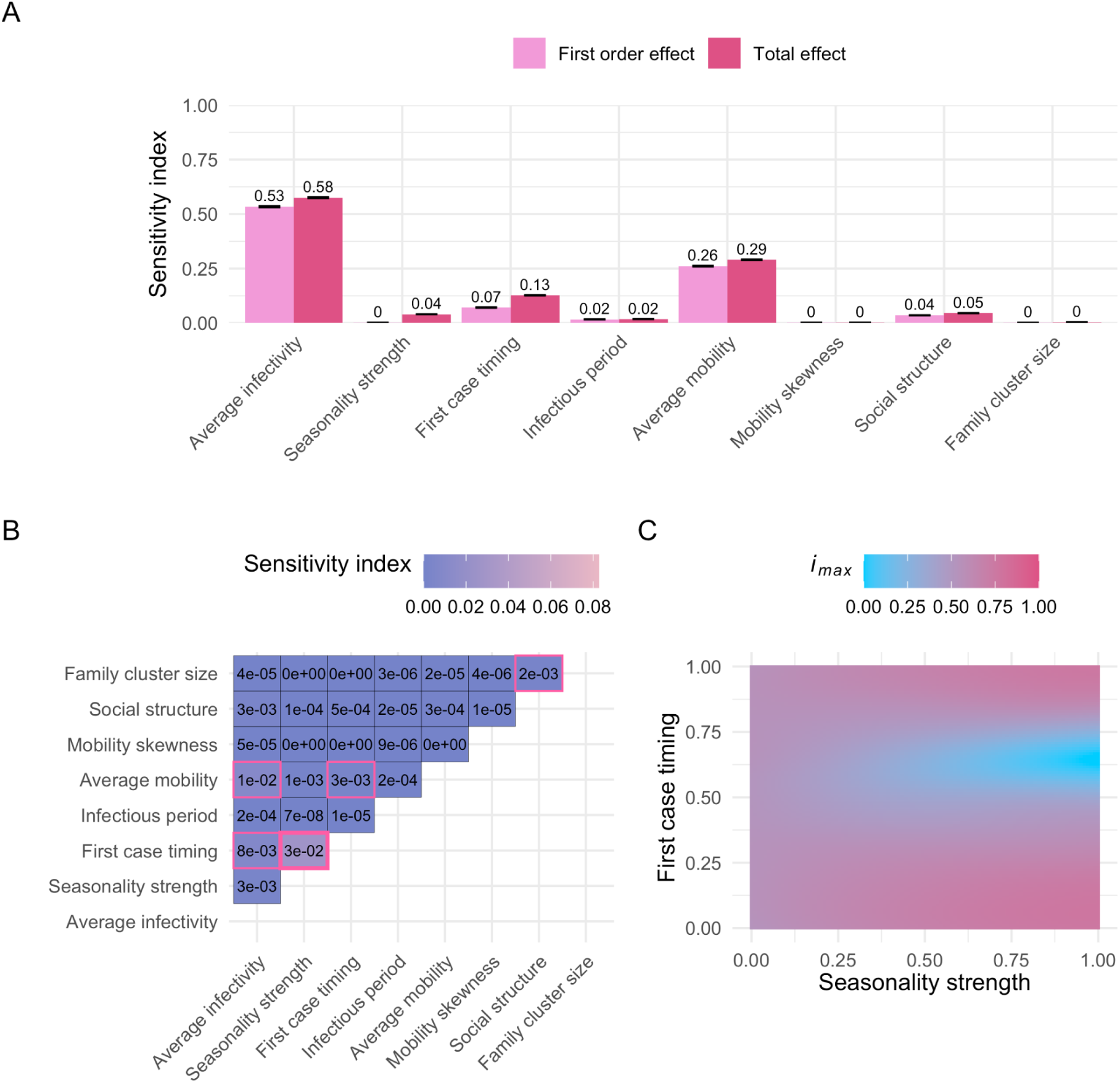
Sobol sensitivity analysis, maximum incidence (*i_max_*). (A) First-order and total effects across the entire input domain (Table 1). The first-order effect describes the impact of a single parameter on the model output (*i_max_*), while the total effect of a parameter accounts for both its first-order effect and all interactions with other parameters. Error bars represent the 95% confidence intervals of the sensitivity index estimates. A total of 9,437,184 points were evaluated for the sensitivity analysis. (B) Second-order effects across the entire input domain (Table 1). A second-order effect captures the pairwise interaction between two parameters. Sobol indices with a 95% confidence interval that does not overlap zero are highlighted with a pink border. The largest second-order effect is emphasized with a bold pink border. (C) *i_max_* predictions with varying “seasonality strength” and “first case timing” parameters (i.e., the two parameters with the largest second-order effect, see panel B). Other parameters were fixed at default values (Table 1). Corresponding sobol sensitivity analysis plots for outbreak probability and outbreak duration can be found in the supplementary figure section (Figures S3 and S4).

The first-order effect estimates for average infectivity are nearly identical across all three metrics: outbreak probability, *i_max_*, and duration (0.52, 0.53, and 0.53, respectively; Figures 3, S3–S4). However, the total effect of average infectivity is notably higher for outbreak probability (0.69; Figure S3) compared to *i_max_* and duration (both 0.58; Figure 3, S4). The higher total-order effect for outbreak probability is primarily driven by the interaction between average infectivity and average mobility (Figure S3B–C). This makes intuitive sense, as highly infectious diseases can still trigger outbreaks even when individual movement is limited. By contrast, the spread of less infectious diseases relies more heavily on sufficient individual mobility to compensate for a lower per-contact infection probability. Accordingly, the first-order effects of average mobility are smaller for outbreak probability compared to *i_max_* and duration (0.12 vs. 0.26 and 0.27). However, the total-order effects of average mobility are similar across all three metrics (0.27, 0.29, and 0.3, respectively, Figures 3, S3–S4).

The reduced importance of the interaction between average infectivity and average mobility for *i_max_*and outbreak duration is due to the fact that these metrics are calculated only across simulations in which an actual outbreak occurred. For these two metrics, the largest second-order effect is the interaction between the seasonality strength and the timing of the first infectious case (Figure 3B, Figure S4B). In our model, the infection probability fluctuates seasonally, following a cosine pattern, and thus the initial infection timing relative to the seasonal cycle is crucial. Introducing the disease during a low-risk season (i.e., when the value of the first case timing parameter falls between 0.25 and 0.75) can lead to prolonged epidemics with lower *i_max_* (Figure 3C, Figure S4C). Since the infection probability depends on the interaction between the average infectivity, the seasonality strength, and the first case timing, it is encouraging that sensitivity analyses of the GP surrogate models effectively uncovered these pairwise interactions (Figure 3B, Figure S3–S4B).

We also observed that family cluster size has only a minor effect on the outcomes, which is mainly driven by interactions with the social structure parameter (Figure 3B, Figure S3–4B). Since individuals return home each day, they are more likely to interact with others within their home location and family cluster (as long as the social structure parameter is > 0). Family cluster size determines how many individuals, on average, live within each family cluster, and in combination with social structure, it influences the extent of interaction within those family clusters, thus having some effect on the investigated model outputs.

As we pointed out earlier, the results of our sensitivity analysis depend on the variance present in the system. When some parameters explain a disproportionately large amount of variance in the model output across the entire input domain, the contribution of other parameters can be hard to detect. To address this, one can fix the former parameters at specific values and then conduct a “conditional” sensitivity analysis that only varies the latter parameters to assess their relative effects in this particular subdomain of the parameter space.

We used this approach to analyze the effect of seasonality strength and first case timing on outbreak probability across various average infectivity and average mobility scenarios, since these parameters were previously identified as crucial drivers (Figures 3, S3–4). Interestingly, we observed a sharp increase in the first-order indices for the first case timing when the average infectivity was fixed at a low value, followed by a gradual decline as the average infectivity increased (Figure 4A). This pattern corresponds to a shift in the system: moving from a state where epidemic outbreaks are rare (Figure 4B: 1^st^ panel) to one where outbreaks occur in the majority of simulated scenarios (Figure 4B: 4^th^ panel). Under conditions that are generally unfavorable for disease transmission, outbreaks are rare and occur only when all parameters align to support an outbreak. Consequently, the first-order sensitivity indices for the first case timing parameter tend to be low, because this metric reflects only the independent effects of single parameters. With increasing average infectivity, the system enters a state where the outbreak probability is primarily driven by the first case timing, creating a hit-or-miss dynamic (Figure 4B: 3^rd^ panel). Here, the strength of seasonality plays a minimal role; the key factor determining whether an outbreak occurs is whether the first case is introduced when infection probabilities are above or below the average infectivity. As average infectivity continues to increase, we observe a U-shape on the outbreak probability heatmap, especially when first case timing coincides with conditions unfavorable for disease transmission (Figure 4B: 4^th^ panel). This reflects how, at this stage, seasonality strength again becomes a critical contributor to outbreak probability. As the average infectivity increases further, the proportion of unlikely outbreak scenarios shrinks, causing the first-order sensitivity index of the first case timing parameter to gradually decrease again. We confirmed that these patterns are not artifacts of the GP model by validating them with the IBM, which showed the same trends (Figure 4C).

**Figure 4.**
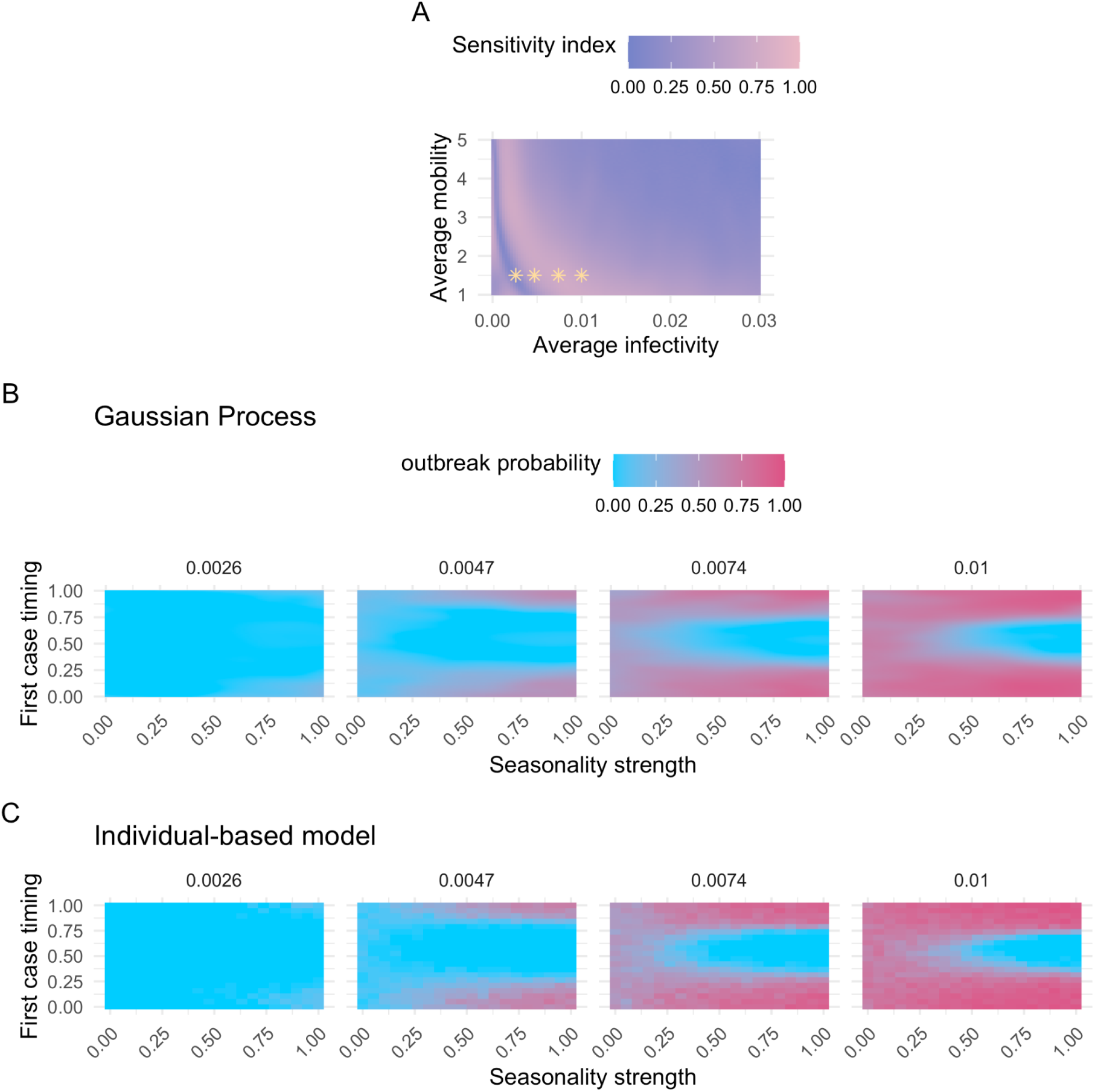
Summary of model outcomes related to outbreak probability. (A) First-order sensitivity index estimates for the first case timing parameter across varying average infectivity and average mobility values. A total of 294,912 points were evaluated for the sensitivity analysis per parameter combination. All other parameters vary across their full ranges (Table 1). The first-order effect measures the influence of a single parameter on the model output (outbreak probability). Yellow stars mark parameter combinations associated with specific model outcomes shown in (B). (B) Predicted outbreak probabilities using the Gaussian Process surrogate model with varying seasonality strength and first case timing values. Panels represent different average infectivities. All other parameters are fixed at default values (Table 1), except for average mobility, which is set to 1.5. (C) Outbreak probabilities inferred from the individual-based model, with varying seasonality strength and first case timing values. Panels represent different average infectivities. As in (B), the remaining parameters are fixed at default values (Table 1), except for average mobility which was set to 1.5. (B) and (C) thus represent model outcomes for the same model parameters, but conducted with the Gaussian Process surrogate model (B) versus the original individual-based model (C), allowing a direct comparison between the two.

### Application to empirical dengue incidence data

To determine whether insights from our sensitivity analysis of the abstract IBM could inform our understanding of real-world epidemics, we analyzed over a decade of weekly dengue incidence data from Colombia [34], along with municipality-level processed demographic and environmental data from Siraj *et al.* (2018) [35]. We retrieved weekly dengue incidence data for Colombia from the OpenDengue database [34], covering January 1^st^, 2007, to December 31^st^, 2019, resulting in 163,279 entries. We selected this cutoff date to avoid confounding effects from the COVID-19 pandemic. To account for potential under-reporting and asymptomatic cases, we adjusted reported dengue incidences by a correction factor of 25 [45,46]. We focused on 211 municipalities with populations of at least 30,000 individuals and dengue maximum incidence rates of at least 0.1%. We defined outbreaks as periods of at least four consecutive weeks during which a smoothing spline fitted to the weekly dengue incidence exceeded the median incidence rate, resulting in the identification of 1,211 epidemic outbreaks.

On this data set, we first tested our model’s prediction of a potential inverse relationship between the average infectivity and average human mobility (Figure S3C). Due to the lack of fine-scale data, we used mosquito abundance probability [35,47] as a proxy for average infectivity. While practical, this simplification does not fully capture the complexity of real-world disease transmission, which is shaped by factors such as local vector control, the level of urbanization, and human-mosquito interactions. For average human mobility, we used the inverse of mean travel time as a proxy [35,48]. Mean travel time is defined as the average time required to reach a settlement with at least 50,000 inhabitants (the settlement might not necessarily be within the municipality itself). We would like to point out that mean travel time reflects regional connectivity rather than fine-scale, within-municipality movement. While this proxy provides some insight into general mobility patterns, it does not necessarily capture the local-scale movement dynamics most relevant to disease transmission. From our model’s predictions, we would expect a positive correlation between mean travel time and mosquito abundance for real-world epidemic outbreaks. However, no significant positive correlation was found, even when restricting the analysis to most remote municipalities (travel time at the 85th percentile or above; Spearman’s rank correlation test: *ρ* = 0.1, *S* = 917,546, *p* = 0.085). This lack of association may partly reflect the limitations of using mean travel time as a proxy for the kind of human mobility that drives local epidemic dynamics.

We then tested our model’s prediction that the timing of the first infectious case should play a crucial role in shaping outbreak dynamics whenever seasonality is important. However, empirical data only allows us to observe outbreaks that have been measurable in the population. We do not know when or how the first infectious case was introduced, nor do we have information about instances where an infectious case was introduced but did not result in an epidemic. When testing across all observed outbreaks, binned by week, we find that the distribution of epidemics is not uniform throughout the year (Chi-squared test: χ^2^ = 117.85, df = 52, *p* < 0.001), which suggests a seasonal effect, and matches our expectations of dengue outbreak dynamics [45,49].

Next, we used the GP predicting *i_max_* to systematically explore whether and how different parameter combinations in our abstract IBM can effectively recapitulate the empirical dengue incidence data from Colombia. To account for heterogeneity in dengue risk across regions, we incorporated municipality-specific average infectivities while keeping other parameters constant across municipalities. We focused on municipalities with at least three epidemic outbreaks, resulting in a dataset of 1,186 epidemics across 173 municipalities. For each municipality, we used 67% of the data for calibration, and withheld 33% for testing. Calibration was done across all municipalities simultaneously. Withholding a portion of the data allowed us to assess the predictive performance of the calibrated GP (which was trained on outcomes of our abstract IBM) in estimating *i_max_* for real-world epidemics.

The best-fitting parameter combination, which minimized the RMSE on the calibration data, revealed an unexpectedly high degree of social structure in our IBM (seasonality strength = 0.16; first case timing = 0.58; infectious period = 4.67; average mobility = 4.39; mobility skewness = 0.47; social structure = 0.99; family cluster size = 4.54; scaling factor = 0.03). However, across the top 1 % (250 out of 25,000) parameter combinations with the lowest RMSE, we observed a wide range of values (Table S1), often spanning the entire parameter range. This suggests that in our IBM, multiple distinct parameter combinations can generate similar epidemic patterns. We also observed variation in average infectivity estimates across municipalities, though certain municipalities consistently exhibited higher average infectivity estimates across the top-ranked (i.e., lowest RMSE) parameter combinations (Figure 5A). Notably, these municipalities showed a notable overlap with previously reported dengue disease clusters [36] and had a significantly higher Gross Cell Product (a measure of economic activity [50]), compared to the remaining municipalities, suggesting a possible link between economic activity and dengue incidence (Figure 5B; *Wilcoxon rank sum test: W* = 1,037, *p* = 0.021).

**Figure 5.**
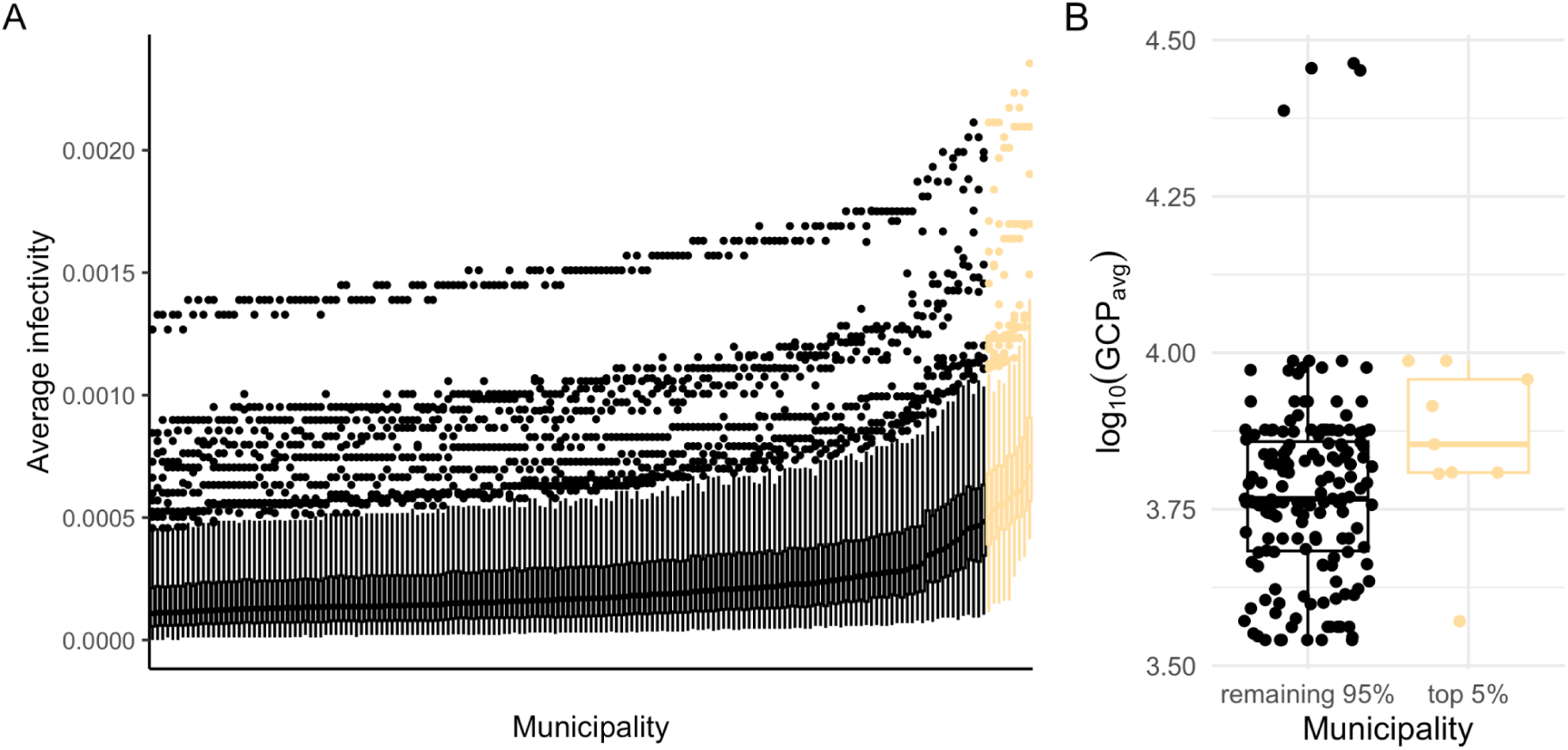
(A) Distribution of municipality-specific average infectivity estimates for the 250 parameter combinations with the lowest root mean square errors. The top 5% of municipalities as sorted by median average infectivity estimates are highlighted in yellow. (B) Average Gross Cell Product (GCP) — a measure of economic activity [50] where higher values represent greater economic activity — distributions, as reported by Siraj *et al.* (2018), for the municipalities depicted in (A), with the municipalities with the largest average infectivity estimates grouped separately.

Finally, we evaluated the predictive power of the GP using the best-fit parameters to predict *i_max_* of epidemics across municipalities for the withheld test data. While the model achieved an RMSE of 0.006, the normalized RMSE (RMSE, scaled by the mean of the observed data) was 1.02, indicating that the model struggled to capture the full complexity of the system and that the model’s predictions were not highly accurate (Figure S5A). The rank correlation coefficient between observed and predicted values was 0.458, with permutation tests placing the model in the top percentile, confirming it has some predictive power. However, these results highlight the limitations of our approach when applied to large-scale, heterogeneous epidemic data.

## Discussion

In this paper, we demonstrated the potential of statistical emulation for studying the dynamics of epidemiological IBMs. Specifically, we implemented an abstract individual-based disease transmission model, loosely inspired by dengue, in C++ and trained Gaussian Process (GP) emulators to approximate three key outbreak metrics: outbreak probability, maximum incidence (*i_max_*), and outbreak duration. Due to their fast prediction speed, these GPs facilitated highly efficient exploration of the model’s eight-dimensional parameter space, allowing us to conduct comprehensive sensitivity analyses that would have been computationally prohibitive using the IBM directly. Our results show that average infectivity and average mobility have large first-order effects and influence all three epidemiological metrics. The most important pairwise parameter interaction varies by model outcome: the interaction between average infectivity and the average human mobility primarily influences outbreak probability, whereas the timing of the first infectious case, combined with seasonality strength, can shape both *i_max_*and the duration of epidemics. Although our trained GP — and the underlying IBM — do not fully capture the full complexity and heterogeneity of real-world dengue dynamics, they provide a computational efficient framework for exploring broad epidemiological patterns and trends. When applied to Colombian dengue incidence data, the approach highlighted municipalities that overlap with previously identified dengue clusters [36], illustrating how statistical emulation can complement empirical research by linking computational modeling with observed disease distributions.

### Individual-based model

The primary aim of this study was to demonstrate the potential of GP emulation as a tool for efficiently analyzing individual-based epidemiological models, rather than to construct a detailed, disease-specific representation of dengue. However, because we applied the framework to Colombian dengue data for illustration, it is important to acknowledge key simplifying assumptions in our IBM relative to known dengue disease transmission characteristics. For example, we modeled an initially completely susceptible population in our simulations, neglecting any preexisting immunities at the onset of the epidemic. Our approach ignores the fact that dengue is caused by four distinct viral serotypes (DENV–1 to DENV–4), and while infection with one strain provides long-lasting immunity against that specific strain, immunity to other strains lasts only a short time [51]. Moreover, a second infection with a different serotype can trigger antibody-dependent enhancement, significantly increasing the risk of severe (and symptomatic) dengue [51]. In hyperendemic countries such as Colombia [45], where multiple dengue virus serotypes are simultaneously circulating within the population, this can cause complex immunity dynamics. Unfortunately, strain-specific sequencing data and antibody measurements that could be used to accurately estimate the proportion of immune individuals are scarce [45].

Furthermore, while our abstract IBM incorporates some key aspects of dengue epidemiology, such as the infectious duration in humans [33] and the role of human movement [37,43], we chose not to explicitly model mosquito vectors. Combining host models with detailed vector models that account for factors such as habitat availability and selection pressures across mosquito life stages could significantly enhance the realism of epidemiological simulations [2,52], albeit at a substantial cost in model complexity, number of parameters, and simulation runtime.

Another simplifying assumption in our IBM is in the human mobility model. While the “family cluster size” and “social structure” parameters allow us to model populations with varying levels of social interconnectivity, locations are not spatially explicit, meaning that the distance between them is not defined. Thus, the likelihood of a person visiting a location is solely determined by parameters affecting social population structure and human mobility. Real-world human movement patterns, on the other hand, are known to exhibit strong spatial regularity [8,53,54]. Moreover, in reality, human populations are rarely closed systems like the one we modeled here. Migration and a variety of factors — economic shifts, environmental changes, large public events — often lead to interactions beyond regular social circles, increasing the risk of disease introduction into areas that were previously unaffected [55].

### Gaussian Processes

While we decided to train our GPs on outbreak probability, *i_max_*, and duration, a GP could instead be trained on other outputs from the IBM. For example, a GP could be trained on the total epidemic size or the time to the epidemic peak, if relevant to addressing the research question at hand. It would also be possible to use a so-called multi-task GP [56], which allows the simultaneous prediction of multiple outputs, and is capable of capturing correlations between them. This could improve the efficiency of the training process, especially when the outputs are highly correlated, because multi-task GPs can leverage shared information between the prediction tasks to enhance accuracy and reduce computational costs. Our choice of separate GPs was guided by two factors. First, the outbreak probability GP was trained on the proportion of simulation runs with observed outbreaks, while the *i_max_* and duration GPs were exclusively trained on simulations with observed outbreaks, which made choosing a consistent set of training points across all three metrics challenging. Second, we had no clear expectations regarding the correlation between *i_max_*and outbreak duration: simulations with shorter durations might result from severe epidemics where most individuals are infected rapidly (high *i_max_*), or from scenarios where the disease quickly dies out (low *i_max_*). These complex dynamics made separate GPs a simpler, more practical choice.

A key factor in implementing a GP is the choice of an appropriate kernel [18,44]. We used the Matérn kernel because of its flexibility in modeling different levels of smoothness in the data. For this kernel, we chose a smoothness parameter *v* = 0.5, which can be beneficial for capturing model behavior in which small changes in parameters can result in abrupt changes in model outputs, as seen here. Preliminary testing, as well as our trained GPs, showed satisfactory performance with the Matérn kernel, so we did not pursue alternative kernels. Whether the accuracy of our GPs could be improved even further with more customized or composite kernels tailored to specific features of the data remains to be explored.

One key advantage of GPs is their Bayesian nature, which allows for uncertainty quantification. This property is particularly useful in active learning, wherein the uncertainty measurements can be leveraged to choose the most informative points to add to the training data. During GP training, we selected half of the new points based on the confidence interval widths, while the other half was selected using the product of the confidence interval widths and a function of the predicted mean. Specifically, we weighted the confidence interval widths based on how close the predicted mean was to its most extreme possible values, assigning the highest weights to intermediate predictions. This approach encourages the GPs to move away from the edges of the parameter space, where uncertainties are naturally higher and predicted means often become extreme. These extremes occur either due to expected model behavior at the parameter boundaries (extreme parameter values cause extreme model behavior), or because data is sparse in these regions, causing the GP to revert to its prior (a constant mean of 0 in our case, which is an extreme value relative to the average predicted value) [18]. However, this approach might overlook regions that the GP does not determine to be highly uncertain but which could provide valuable information if explored. Alternative sampling strategies, such as expected improvement, could help identify points that boost model performance, even if their initial uncertainty is lower. Moreover, tools like BoTorch [57] provide libraries to implement advanced batch optimization techniques, allowing the selection of sets of data points that are chosen together to maximize their combined impact on improving GP performance. While more advanced techniques like expected improvement scores and batch optimization could potentially enhance GP performance, they would require further model tuning and validation, which is beyond the scope of this study.

### Sensitivity analysis

The fast prediction speed of the trained GPs allowed us to conduct comprehensive variance-based sensitivity analyses. However, this approach could be confounded by potential discrepancies between the GP surrogate model and the original IBM. While the GPs generally predicted epidemiological metrics inferred from the IBM well, the width of the sensitivity analysis confidence intervals should be interpreted cautiously. Furthermore, average infectivity and average mobility emerged as the dominant contributors to variance in the epidemiological metrics, making it harder to detect the influence of the other parameters. This scaling effect can obscure smaller, but still relevant, factors. To address this, we also performed sensitivity analyses in targeted regions of the parameter space for which average infectivity and average mobility were fixed, revealing state changes within the model’s dynamics — sudden transitions from rare epidemic outbreaks to frequent outbreaks — which were confirmed by simulating selected points directly with the IBM. However, it is important to note that while a sensitivity analysis captures the variance in model outputs due to parameter changes, it does not fully capture the underlying dynamics of the model, such as state transitions or the mechanistic interactions between single parameters that drive these changes. Specifically, the sensitivity analysis highlights which parameters contribute most to the output variance, but it does not reveal why certain parameter combinations lead to changes in the model behavior.

We observed the largest second-order effects between average infectivity and average mobility for the outbreak probability metric, and between seasonality strength and first case timing for *i_max_* and duration. To explore how well these model-derived insights translate to real-world epidemic outbreaks, we examined over a decade of dengue incidence data from Colombia. We used mosquito abundance probability [35,47] as proxy for average infectivity, implicitly assuming a constant biting rate whereby higher mosquito abundance directly translates to increased infection probability. However, this simplified representation overlooks the complexity of real-world disease transmission dynamics, which is shaped by factors such as vector control [58], urbanization, human-mosquito contact rates [59], and mosquito behavior [60]. We did not observe the expected correlation between our proxies for average human mobility and average infectivity in the empirical outbreak data. This may be partly due to previous findings that mean travel time serves as a broad indicator of accessibility rather than a precise measure of actual human mobility [35]. While our analysis supports a seasonal pattern of dengue outbreaks consistent with prior studies [45,49], we treated each epidemic as independent, not accounting for temporal correlations within municipalities or spatial dependencies across neighboring or well-connected municipalities. As a result, the statistical significance of our findings should be interpreted cautiously. Overall, these results illustrate both the potential and the current limitations of applying abstract individual-based model insights — based on a simplified disease transmission framework — to empirical epidemiological data.

### Application to empirical dengue incidence data

When comparing the predictions of our model with real-world outbreaks, the only empirical parameter that actually varied between epidemics within each municipality was the epidemic’s onset. This limited the GP’s flexibility to generate diverse predictions within each municipality. In fact, a simple linear mixed-effects model that predicts log_10_-transformed *i_max_* values based on the onset timing of an epidemic, while accounting for the municipality-level variations with random effects, performed similarly to the GP model on withheld test data (Spearman’s *ρ* = 0.54). This suggests that both the abstract IBM and the GP emulators might be too generalized to effectively predict real-world outbreak data across multiple municipalities. To achieve more accurate predictions, the IBM would need to be more complex, incorporating municipality- and disease-specific characteristics such as outbreak histories, population immunity levels, and finer-scale human movement patterns, which might be critical for capturing the nuanced dynamics of local outbreaks.

Despite the GP emulator’s (and the underlying IBM’s) limitations in capturing the full complexity of empirical dengue dynamics, our analysis revealed that a subset of municipalities consistently exhibited higher average infectivity estimates. Several of these municipalities — such as Puerto López, Leticia, Melgar, and La Mesa — stand out due to their economic or geographic context. For example, Puerto López, which had the highest average infectivity estimate, is a key river port, while Leticia is located at the tri-border area of Colombia, Brazil, and Peru, functioning as a major hub on the Amazon river. Tourist destinations such as Melgar and La Mesa also showed elevated average infectivity estimates, potentially reflecting increased human movement and connectivity driven by tourism and travel — factors that may enhance dengue transmission in these areas. These observations support the idea that human movement and economic activity could play a significant role in shaping dengue dynamics [61].

At the same time, municipalities with higher average infectivity estimates also tended to have greater economic activity, which may be associated with better healthcare access and, in turn, increased detection and reporting of dengue cases. This introduces potential bias, because our model assumes constant reporting rates across municipalities, highlighting the need for caution when interpreting these findings. Nonetheless, many of the municipalities with elevated average infectivity estimates are located in Colombia’s western regions, overlapping with areas previously identified as disease clusters for dengue and other *Aedes*-borne diseases [36].

## Conclusion

In conclusion, we explored the utility of statistical emulation to efficiently analyze epidemiological IBMs. The use of GP-based emulators allowed valuable insights into the key drivers of our simulated disease dynamics, revealing critical interactions between average infectivity, human mobility, and seasonality. Overall, our work demonstrates both the potential and the challenges of using statistical emulation to explore complex epidemiological systems, providing a foundation for future efforts that could incorporate additional model complexity and realism while maintaining computational efficiency.

## Methods

### Individual-based model

This detailed individual-based model (IBM) description is motivated by previous studies using the ODD protocol for describing IBMs [1,5,62]. The IBM was implemented in C++.

#### 1. Purpose and patterns

The purpose of our IBM is to explore how human movement, social population structure, and seasonal variation in infection probability influence infectious disease dynamics. Rather than replicating a specific empirical system, the model is intended to highlight the relative importance of these parameters and their interactions in shaping disease spread. Its performance is evaluated by examining how the absolute number of infectious cases and the change in infectious cases over time are affected by IBM parameter changes.

#### 2. Entities, state variables, and scales

The model operates in daily timesteps and includes two primary entities: humans and locations. Each location represents a residential home with a group of residents, who are collectively considered a ‘family’. There are no distinct non-home locations such as workplaces, schools, or other public spaces. The number of humans in a given family — the number of humans residing at a given home location — is drawn from a negative binomial distribution with *μ* = 6.2 *and θ* = 9.07, following the household size distribution observed in Iquitos, Peru [37,40].

Each individual human has the following state variables: a home location, an infection status (susceptible, exposed, infected, or recovered), the number of remaining days in their current infection status (how much longer they will stay in their current state), and the number of elapsed days in their current infection status (how long they have already been in that state).

The basic spatial unit of the model is the location. Simulations are initialized by generating 10,000 locations. Each location has the following state variables: the number of infectious humans visiting it in the current timestep of the model, its per-contact infection probability (the probability that an infectious individual that visited the location at time *t-1* transmits the disease to a susceptible individual that visits the same location at time *t*), a history of the number of infectious visitors it has had, and a history of its infection probability over time. All locations are randomly grouped into family clusters of a user-specified size. Each location is assigned to only one family cluster, ensuring that no locations are shared between clusters. All family members of a single home location belong to the same family cluster, but not all members of a given family cluster reside in the same home location. Figure S1 provides a conceptual overview of family clusters (and human movement) as implemented in our IBM Conceptually, then, a family cluster is a group of locations, the members of which tend to socialize together; an individual in a given family cluster is more likely to visit a location inside the family cluster than to visit a location outside it. This grouping introduces social structure into the simulation (please refer to the next section for further details). Note that “family clusters” are conceptually equivalent to the “social groups” defined by Reiner *et al.* (2014) [40].

#### 3. Process overview and scheduling

The two core processes of the model are human movement and infection dynamics.

##### Human movement

For the process of human movement, during each timestep the model iterates through all individuals to determine which locations they will visit. Each individual visits its home location at least once every timestep. For each individual, an additional number of visits by the individual is drawn from a negative binomial distribution at the beginning of each day. The use of a negative binomial distribution allows for heavy-tailed human mobility distributions where some individuals are highly mobile, visiting a large number of locations per day. However, there is no correlation between the various sampled values for a given individual over time, meaning that we did not model systematic super-spreaders in our IBM. The infection status does not affect human movement in our IBM: infectious individuals visit, on average, the same number of locations as susceptible or recovered individuals.

For each visit, whether the visit is to a location inside the individual’s family cluster or to a location outside that cluster is determined probabilistically, with the social structure parameter being the probability that a particular visit happens within the family cluster (Table 1). Locations to be visited are then randomly selected from the set of locations inside or outside the individual’s family cluster, as appropriate. Multiple visits to the same location are allowed. Figure S1 provides a conceptual overview of human movement patterns as implemented in our IBM.

##### Infection dynamics

In our model, each human can have one of four infectious states: susceptible, exposed, infectious, and recovered. The model tracks the number of days each simulated human will remain in its current infection state, decreasing this count at the end of each day.

At the start of the simulation, all humans are susceptible. The disease is then introduced into the population by randomly selecting one individual and immediately changing their infection status from susceptible to exposed. This exposed status indicates that the individual has contracted the disease but is not yet infectious. Unless specified otherwise, exposed individuals become infectious at the end of each day, effectively reducing the model to a Susceptible-Infectious-Recovered model, where the length of the infectious period is specified by the user. At the end of an individual’s infectious period, the individual’s infection status changes to recovered the next day. The model assumes lasting immunity, so once individuals recover, they cannot be reinfected.

If a susceptible human visits a location that had *N* infectious visitors the day before, the probability of contracting the disease and immediately entering the exposed state is 1 − (1 − *p_infection_*)^*N*^ where *p_infection_* is the infection probability per contact. (This is simply the probability that a binomial draw B(*N*, *p_infection_* (*t*)) ≥ 1, indicating that infection occurred from at least one previous visitor.) The infection probability *p_infection_* follows a cosine function that is determined by three parameters: the average infectivity (α_0_), the seasonality strength (α*_season_*), and the first case timing (*t_first_*), and is calculated as follows:

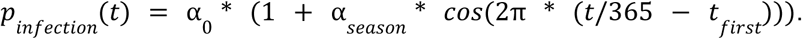

Each location is assigned the same infection probability for a given day, as determined by the cosine function above. Variations in the overall likelihood of infection, from location to location, arise from the differing numbers of infectious individuals visiting each location.

If a visiting human is already infectious, the model increments the count of infectious visits for the current day at each location visited by that human, which will make those locations “infectious” in the following timestep as just described.

Since humans only change their infection status at the end of each day, and the likelihood of infection for susceptible individuals is determined by the number of infectious visitors from the previous day, the order in which individuals are processed is inconsequential. This ensures that the model remains asynchronous and order-independent during each day. Indeed, this design would allow the model to be parallelized to run across multiple processing cores, although runtimes were fast enough that we did not deem that necessary.

#### 4. Design concepts

##### Basic principles

The model aims to study abstract disease dynamics within human populations exhibiting varying levels of social structure. It does not focus on the realistic modeling of a specific city, or on the biological details of a specific disease.

##### Emergence

The number of infectious individuals each day — the central output of this model, for our purposes — is an emergent property, not predefined within the model. Stochasticity plays a major role in introducing uncertainty into these patterns.

##### Adaptation, objectives, learning, prediction, and sensing

None of the individuals in the model have the ability to adjust their behaviors. There are no adaptive behaviors, learning abilities, predictive capabilities, or sensing capabilities in the model.

##### Interaction

Humans interact by potentially infecting other humans who visit the same location the next day.

##### Stochasticity

This IBM incorporates stochasticity in the family sizes, the daily number of visits per human, and the probabilistic infection dynamics. For detailed descriptions, please refer to Sections 2 and 3.

##### Collectives

Each human is assigned a home location and a family cluster, making them members of a family and a collection of locations. However, no special properties are attributed to sharing a home location or family cluster, except for the general tendency to interact more frequently due to the human movement rules described in Section 3.

##### Observations

The model outputs a table of the counts of susceptible, exposed, infectious, and recovered individuals for each day of the simulation, across the entire population.

#### 5. Initialization

Please refer to Sections 2 and 3 regarding the initialization of the model.

#### 6. Input

The model has eight parameters that can be specified by the user with command-line arguments (Table 1).

Out of these eight parameters, three parameters collectively influence the infection probability, as described by the equation in Section 3: the average infectivity, the seasonality strength, and the first case timing. Additionally, the user must define:

- The length of the infectious period, after which infectious individuals transition to being recovered (Section 3: *infection dynamics*).
- Parameters for the negative binomial distribution describing human movement (Section 3: *human movement*).
- The proportion of visits that occur within the family cluster of an individual (Section 3: *human movement*).
- The number of locations per family cluster (Section 2).

### Gaussian Processes

Statistical emulation involves replacing an individual-based simulation framework with a statistical surrogate model, or machine learning model trained on input-output pairs from the original framework [12]. For this task, we used GPs, which are non-parametric models that define a distribution over functions. This allows GPs to efficiently interpolate between scarcely sampled observations. GPs are an attractive choice, not only due to their mathematical tractability, but also because their Bayesian nature allows for the quantification of model uncertainty. We incorporated these uncertainty estimates into our policy for scoring potential additional data points (Figure 1A). We implemented GPs in Python (v3.10.6) using the GPyTorch library (v1.11) [19] for efficient GP modeling, and the torch.cuda module from the PyTorch package (v2.0.1) [63] for GPU acceleration with NVIDIA GPUs. The implementation was inspired by a GP surrogate model previously used to study the efficiency of gene drives in rat populations [16].

We trained a separate GP model for each of the following three outbreak metrics: outbreak probability (proportion of simulation runs in which more than 0.1% of the population becomes infected), maximum incidence *i_max_* (highest proportion of infectious individuals at any day), and epidemic duration (timespan from the first infectious case to the recovery of the last infectious individual). While the IBM outputs for outbreak probability and *i_max_*are bounded between 0 and 1, epidemic duration spans a much wider range: In the initial training dataset (N = 5,000 data points), the observed duration of epidemics ranged from 19.65 to 424.15 days. To manage the variance in epidemic duration and improve the GP’s ability to predict longer epidemics, we applied a logarithmic transformation to the outbreak duration.

The response variable for each GP is calculated from 100 simulation runs per parameter combination. While outbreak probability is defined as the proportion of simulation runs with observed outbreaks, *i_max_*and outbreak duration are exclusively calculated from simulations with epidemic outbreaks. This approach helps minimize unrepresentative input-output pairs. For example, if a specific parameter combination results in an *i_max_* of 10%, but only 50% of the simulation runs result in an actual outbreak, averaging across all simulations would result in an *i_max_* estimate of about 5%, not accurately representing the dynamics of the IBM.

The covariance function — or kernel — of a GP determines how much the response values of different input points covary [44]. Thus, the choice of kernel is crucial in shaping the GP’s predictions. We selected a Matérn kernel with *v* = 0.5, which corresponds to the exponential kernel. This kernel is capable of capturing abrupt changes in function values [18]. We applied the same kernel type across all three GPs.

### Gaussian Process training loop

One key advantage of GPs over other machine learning methods is their Bayesian nature, which allows us to identify regions in the parameter space where the model’s predictions are highly uncertain. This allows us to strategically select new training data from areas where the GP is least confident, thereby enhancing prediction accuracy. This approach forms the basis of the GP’s active learning loop (Figure 1A), which consists of three steps: training the GP, scoring potential new training data, and then updating the training data with selected new data points [18].

#### Step 1: GP training

We trained each GP for 16 rounds: one initial round with a training dataset consisting of a Latin hypercube sample (LHS) of 5,000 data points from the entire input domain (Table 1), followed by 15 active training rounds (Figure 1A). For training, we utilized the Adam optimizer from PyTorch [63] with a learning rate of 0.01. In each training round, the GP was trained for 30,000 iterations, with a model snapshot saved every 1,000 iterations. To avoid overfitting, we evaluated all 30 snapshots against a separate validation dataset consisting of 10,000 LHS points. We selected the snapshot with the lowest RMSE on the validation dataset for step 2 in the training loop.

#### Step 2: Data scoring

In each active learning round, we scored 10^7^ LHS points using two distinct policies [16]. These scores are used as probability weights to select 1,000 new data points to expand the training data. Policy 1 is based solely on model uncertainty. In this policy, the probability *p_i_* that a data point *i* is selected is proportional to the width of the 95% confidence interval for that point (*w_i_*), normalized by the total width of all potential data points:

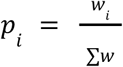

Policy 1 assigns larger weights to data points with greater uncertainties. However, regions with large uncertainties are often clustered near the edges of the observed parameter space, where the GP must extrapolate far beyond observed training data [44]. However, while the uncertainty bounds of these points might be relatively high, the degree of improvement the GP can gain from sampling points from the edges of the parameter space can be limited. To avoid oversampling these areas, we developed policy 2. Policy 2 reduces the likelihood of sampling points with extreme predicted values. Specifically, the 95% confidence intervals from policy 1 are further weighted by the GP’s prediction. The probability *p_i_*of selecting a data point *i* is given by:

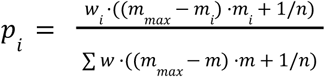

where:

- *w_i_* is the 95% confidence interval width for point *i*
- *m_i_* is the GP’s predicted value for point *i*
- *m_max_* is the maximum predicted value (3 for duration, 1 otherwise)
- *n* is the total number of potential data points

This formulation ensures that points with high uncertainty yet with predicted values near the midpoint of the range are assigned the highest weights. The GP’s predictions were clipped to the range [0, 1] for outbreak probability and *i_max_*, and to [0, 3] for epidemic duration (since the GP predicts log_10_-transformed durations, this range corresponds to durations between 1 and 1,000 days).

For our adaptive sampling strategy, 50% of the points are selected using policy 1, and the remaining 50% are selected using policy 2.

#### Step 3: Update training data

As mentioned earlier, the data points for *i_max_* and duration are based solely on simulation runs where epidemic outbreaks occurred. If a selected data point did not result in 100 outbreaks after 2,000 simulation attempts, we chose a new data point. For the initial training dataset, where no GP predictions were available, this selection was done randomly from 10^7^ LHS samples. In the active learning rounds, we chose all of the new data points as described in step 2. After successfully simulating all selected points, the new results are added to the training dataset, and a new GP training cycle begins (Figure 1A).

### Gaussian Process usage

We evaluated the accuracy of the trained GP using an independent test dataset of 10,000 LHS points and calculated the RMSE. For visualization purposes, such as in heatmaps (e.g., Figure 3C), we clipped the predictions to the range [0, 1] for epidemic probability and *i_max_*, and to [0, 3] for the log_10_-transformed epidemic duration.

### Sensitivity analysis

To explore how changes in parameters affect the epidemic metrics, we conducted a series of variance-based sensitivity analyses [64] in Python using the Sobol method implemented in the SALib library (v1.4.7) [65]. These analyses were performed using the GP’s predictions rather than IBM output. This choice was driven by the substantial computational cost of running the IBM at the scale required for sensitivity analysis: the number of simulations needed for robust estimation of Sobol indices is large [65], and achieving narrow confidence intervals would be computationally prohibitive with the IBM alone. In contrast, the GP emulator offers near-instantaneous predictions and thus enables efficient and tractable exploration of the model’s eight-dimensional parameter space.

The Sobol method quantifies the contribution of single parameters and their interactions to the variance of a model’s output. Since these variance components are often not analytically tractable, the Sobol method approximates them using a Monte Carlo method. The resulting sensitivity indices — first, second, and total order — provide a measure of each parameter’s influence. First-order effects measure single parameter contribution, second-order effects measure the interactions of two parameters, and total order effects capture the combined impact of each parameter, including all interactions with other parameters of any order.

To perform the sensitivity analysis, the number of model evaluations is proportional to *n * (2d + 2)*, where *n* is the base sample size and *d* is the dimensionality of the parameter space (*d* = 8; Table 1) [65]. The accuracy of the Sobol indices improves with a larger *n*, leading to smaller confidence intervals. For the sensitivity analysis of the entire input domain, where all parameters vary across their full range (Table 1, Figure 3A–B), we selected *n* = 2^19^. To investigate the first-order effect of the first case timing with the two most influential parameters (average infectivity and average mobility) held constant, we conducted a sensitivity analysis with *n* = 2^14^ for each combination of these parameters (Figure 4A). We calculated 95% confidence intervals of the Sobol indices using the bootstrapping method provided by SALib [65].

### Empirical data

We retrieved weekly dengue incidence data at the municipality level for Colombia from the OpenDengue database, an open-access platform that provides detailed epidemiological data on dengue [34]. The selected dataset spans from January 1^st^, 2007, when weekly resolution data became consistently available, to December 31^st^, 2019, comprising 163,279 entries. We chose to end in 2019 to avoid the potential confounding effects of the COVID-19 pandemic [66]. We chose Colombia for this study because it is one of the countries most affected by dengue in the Americas [45] and offers exceptionally well-documented time series data on dengue incidence [34]. To adjust dengue incidences for potential under-reporting and asymptomatic cases, reported dengue incidences were corrected by a factor of 25 [45,46]. Estimates for under-reporting factors in dengue typically range from 10 to 27, depending on the region and study [46]. While we recognize that using a correction factor of 25 for Colombia, which has well-documented dengue incidence records [34], might be cautious, we chose it to include a broad range of municipalities.

We obtained municipality-level processed data from Siraj *et al.* (2018), which provides a global, high-resolution dataset of potential environmental drivers for Zika transmission in Colombia between January 1^st^, 2014 and October 1^st^, 2016. Although the data published by Siraj *et al.* (2018) focused on Zika, it is relevant to dengue because both viruses share a primary vector, *Ae. aegypti*, which is responsible for the majority of dengue transmission in Colombia [67]. Specifically, we used four metrics from Siraj *et. al* (2018) (i) the population count (ii) the weekly occurrence probabilities of *Ae. aegypti* [47] (iii) the Gross Cell Product, which measures economic activity at a fine spatial scale [50], and (iv) the mean travel time to the nearest city. Please refer to Table 1 in Siraj *et al.* (2018) for additional information on the municipality-specific data.

We matched the records from Siraj *et al.* (2018) and Clarke *et al.* (2024) based on the names of municipalities and their respective departments. To improve the matching, we standardized the municipality and department names by converting them to lowercase and applying a latin-ascii transformation to remove any accents or special characters. In cases where mismatches occurred, either at the municipality or departmental level, we followed a similar approach to Clarke *et al.* (2024), manually reviewing the records and checking the geographic boundaries using shapefiles. While we were able to obtain the original shapefiles from Clarke *et al.* (2024) (Oliver Brady, personal communication), the original shapefiles for Siraj *et al.* (2018) were not accessible at the time of our study. As a substitute, we used shapefiles from the OCHA database [68]. Despite this limitation, we successfully matched 95% (1,009 out of 1,063) of the municipalities present in the raw dengue incidence data. Our goal was not to achieve a perfect match, but rather to secure a sufficient number of high-quality matches to proceed with our analysis.

We focused on 211 municipalities that aligned with our IBM in terms of population size (i.e., at least 30,000 individuals) and had a maximum dengue incidence rate of at least 0.1% over the entire study period. To detect epidemics, we fitted a smoothing spline using the ss() function from the npreg R-package (λ =10^-10^) [69]. An epidemic outbreak was defined as a period of at least four consecutive weeks in which the spline function exceeded the median dengue incidence rate. Using this method, we identified 1,211 potential epidemic outbreaks with an *i_max_* of at least 0.1% which were included in the analysis. On average, each municipality had 6.34 outbreaks. The average duration per outbreak was 195 days, with an average *i_max_* of 0.6%.

### Parameter exploration with Gaussian Processes

The speed of the GP predictions allows for an efficient exploration of which parameter combinations in our epidemiological IBM provide the best alignment between predicted simulation model output and the empirical dengue incidence data from Colombia. This also allows us to assess whether our GPs — and consequently, our IBM — can provide insights into real-world epidemic outbreaks. To capture the heterogeneity in dengue transmission potential, we incorporated municipality-specific average infectivities while assuming that other model parameters remained constant across all municipalities. We focused on municipalities with at least three outbreaks, resulting in a total of 1,186 epidemics across 173 municipalities. For each municipality, we randomly split the data into 67% for parameter calibration (N = 737) and 33% for testing (N = 449), using the test data to evaluate the GP’s predictive performance for *i_max_*.

We generated 25,000 LHS parameter combinations from the full parameter space (Table 1), excluding the average infectivity parameter. Additionally, we introduced a scaling parameter (ranging from 0 to 0.1) to account for differences in incidence magnitude between the simulated and empirical data. This adjustment was necessary because the GP emulators were trained on IBM model outputs at daily resolution, whereas the available dengue incidence data are aggregated weekly. Since the comparison with empirical data was developed after emulator training, rescaling allowed us to align the model outputs with the empirical data range without retraining. Importantly, the scaling was applied uniformly across all incidence values, preserving relative differences between outbreaks and ensuring comparability across municipalities. For each of the 25,000 LHS samples, we tested 50 evenly spaced average infectivities over the range [0, 0.03], resulting in 1.25 million predicted data points for the 737 epidemics used for calibration. For each epidemic, we accounted for the epidemic’s start time by adjusting the first case timing parameter, while using municipality-specific infection probabilities. The predictions were constrained to the range [0, 1] before calculating the RMSE between observed and predicted *i_max_*. We then selected the average infectivity that minimized the RMSE for each of the 25,000 LHS in each municipality. The 25,000 parameter combinations were ranked by summing the RMSE across all municipalities, using the best-fit (i.e., lowest RMSE) infection probabilities for each. To further investigate infection probabilities across municipalities, we examined the 250 LHS combinations with the lowest RMSE sums.

Finally, we evaluated the GP’s predictive performance on the withheld test data by calculating both the RMSE and Spearman’s rank correlation coefficient (*ρ*). To test the significance of Spearman’s *ρ*, we conducted 1,000 permutation tests, where the start time and municipality for each epidemic in the test set were randomly shuffled.

### Statistical analysis

Unless stated otherwise, statistical analyses were performed using the R statistical computing environment (v4.2.1) [70]. Significance is declared at an alpha cut-off of 5%.

## Code availability

The source code of the individual-based disease transmission model implemented in C++ is available on GitHub at https://github.com/AnnaMariaL/DengueSim. Simulated data, pre-trained GPs, and Jupyter notebooks demonstrating the usage of the GPs are also available on GitHub at https://github.com/AnnaMariaL/DengueSim-GP.

## Acknowledgements

We thank all members of the Messer and Murdock lab for helpful discussions. Special thanks to Beliz Erdogmus for her contributions during the early phases of the project; Isabel Kim, Mitchell Lokey, and Meera Chotai for technical support; Amir Siraj for support with the municipality-specific environmental data; and Oliver Brady for providing supplemental shape files. This project has received funding from the European Union’s Horizon2020 research and innovation program under the Marie Sklodowska-Curie grant agreement No. 101025586. PWM was supported by the National Institutes of Health under award R35GM152242.

## Glossary

GP: Gaussian Process
IBM: Individual-based model
RMSE: Root mean square error
LHS: Latin hypercube sample
*i_max_*: Maximum incidence

## Supplementary information

### Supplementary figures

**Figure S1.**
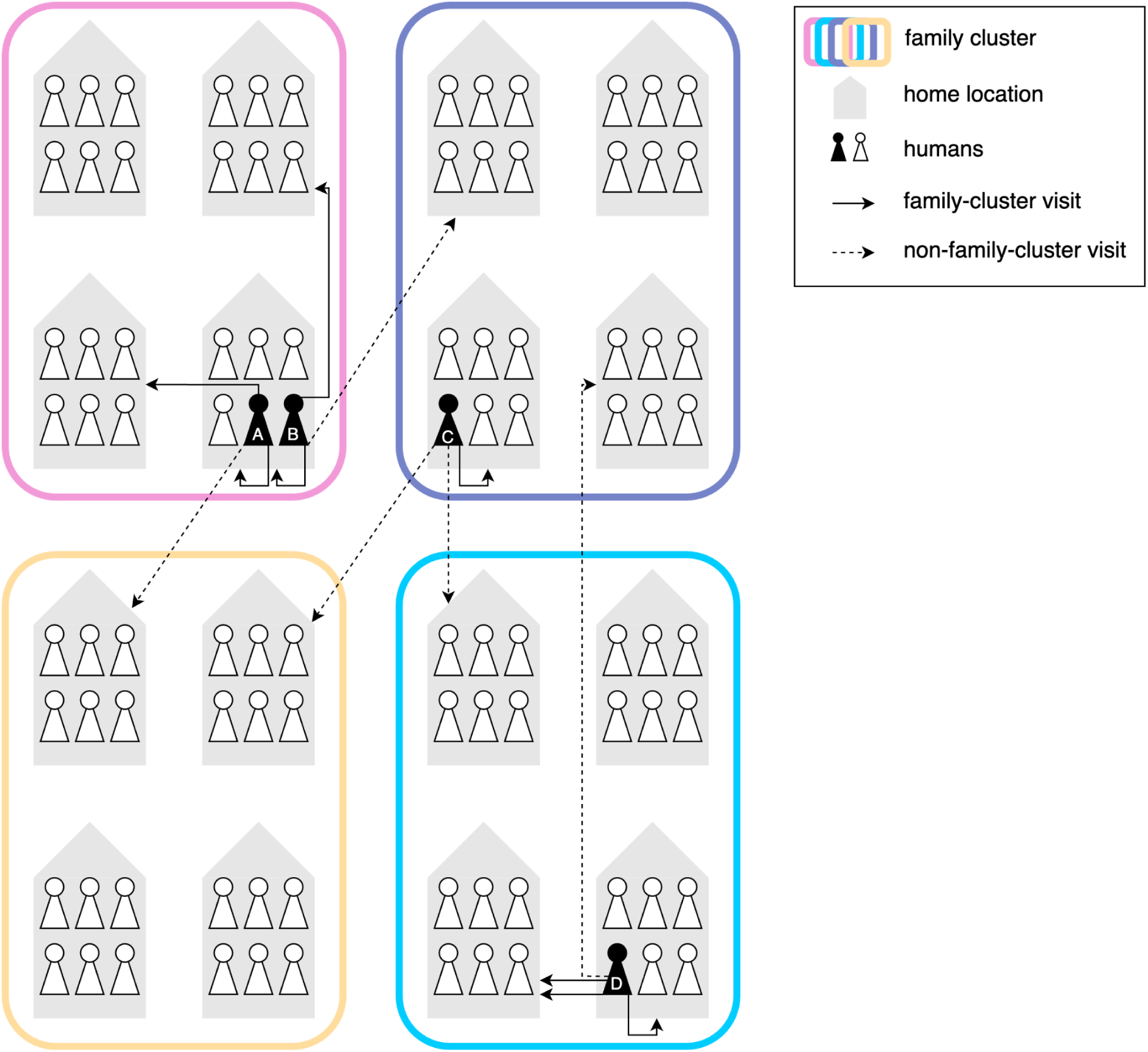
Schematic overview of human movement in the individual-based model. Each colored frame represents a unique, non-overlapping family cluster, with each cluster containing multiple family homes. Individuals can make visits within their own family cluster (solid arrows) or to other clusters (dashed arrows). The likelihood of visits occurring inside the family cluster is determined by the social structure parameter (Table 1). Each individual visits their home at least once per day and moves independently of others in the same family (individuals A and B). Multiple visits to the same location are allowed (individual D). Visits to other family clusters occur randomly and are not restricted to any specific cluster (individual C).

**Figure S2.**
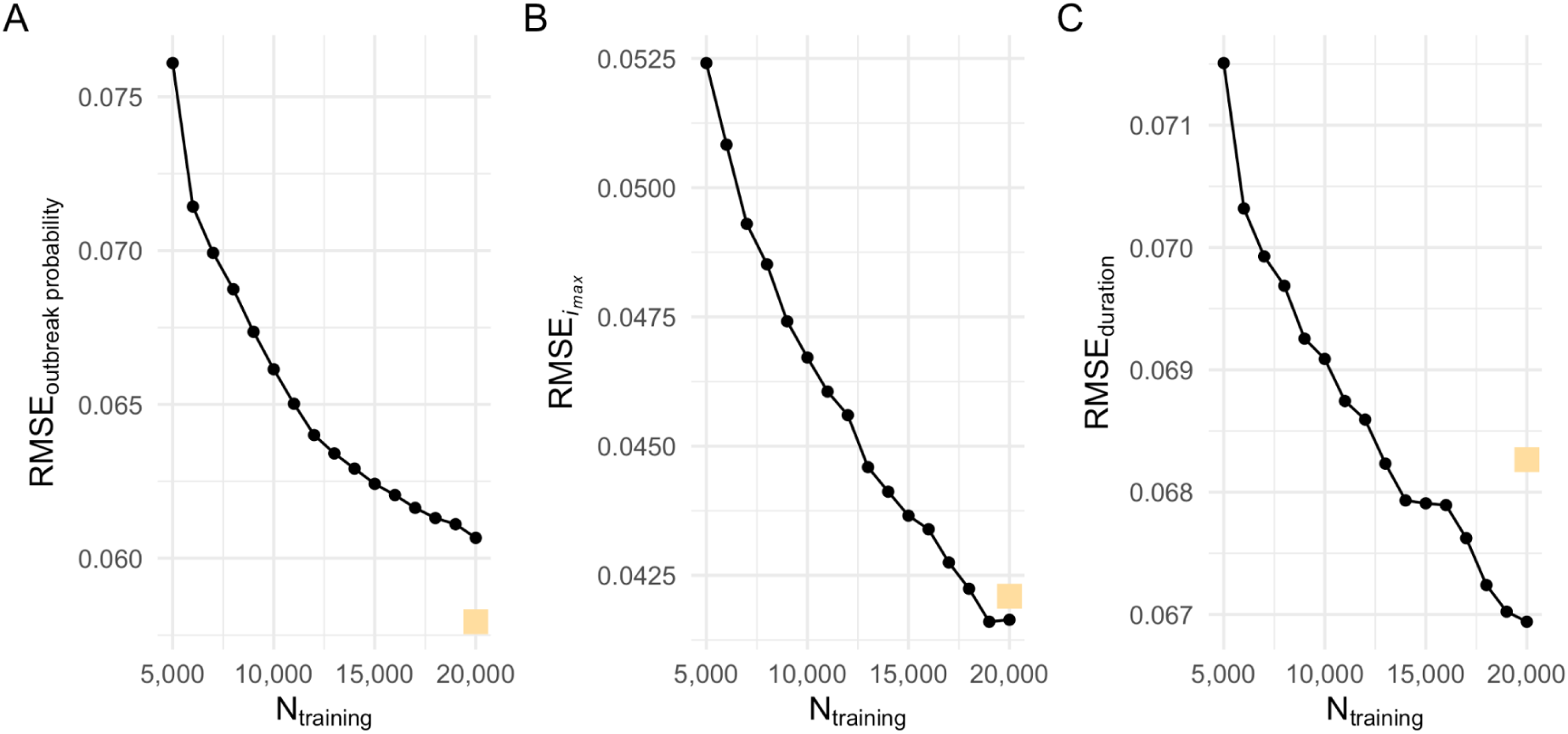
The Root Mean Square Error (RMSE) between the Gaussian Process predictions and the individual-based model results in the validation dataset (N = 10,000 data points). The RMSE decreased as the size of the dataset used to train the Gaussian Processes increased (x-axis). The RMSE between the predictions of the final GP model and the test data (N = 10,000 data points) is indicated by a yellow square. (A) outbreak probability (B) maximum incidence (*i_max_*), (C) log_10_-transformed duration.

**Figure S3.**
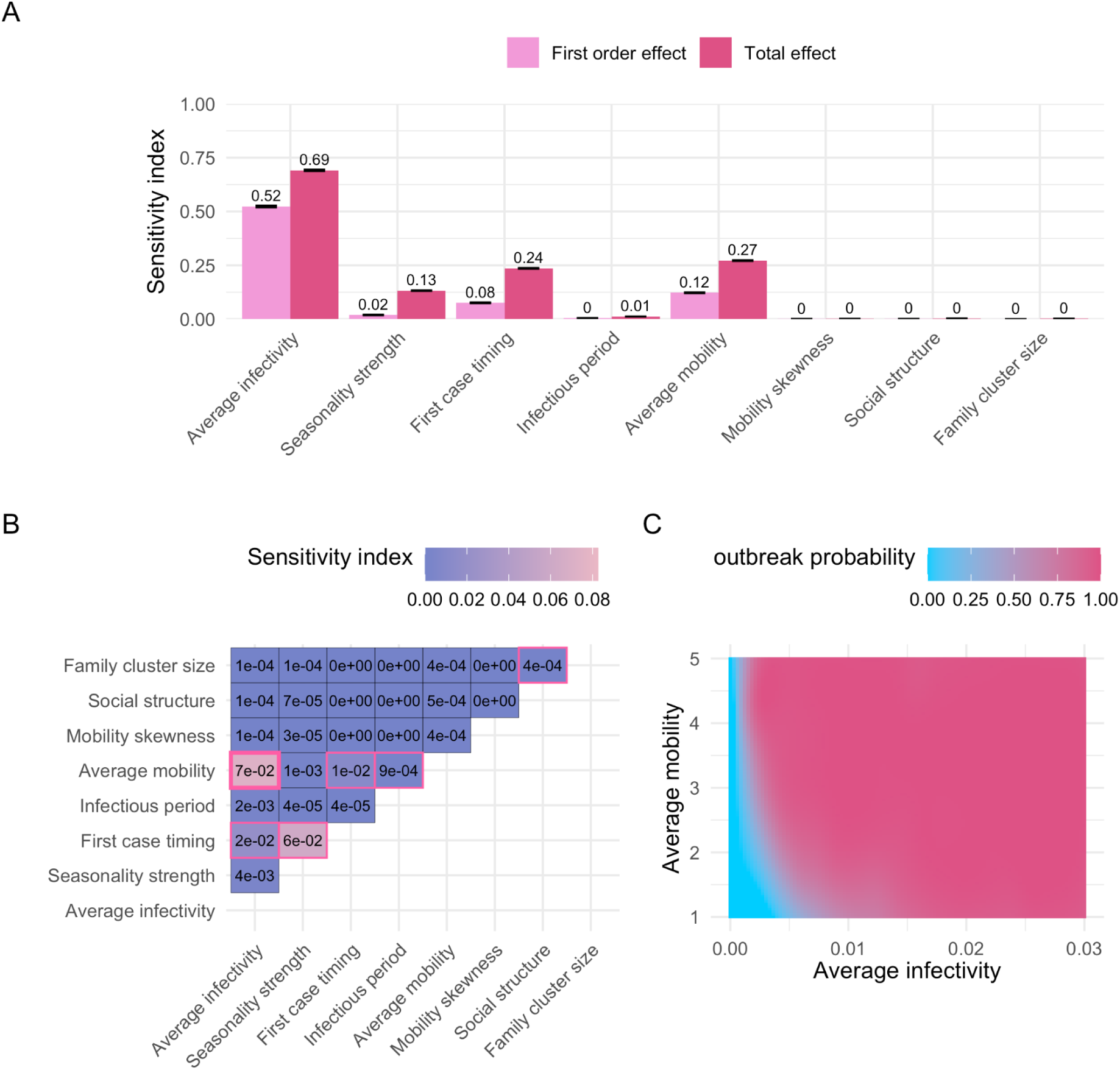
Sobol sensitivity analysis, outbreak probability. (A) First-order and total effects across the entire input domain (Table 1). The first-order effect describes the impact of a single parameter on the model output (outbreak probability), while the total effect accounts for all interactions involving one or more parameters. Error bars represent the 95% confidence intervals of the sensitivity index estimates. A total of 9,437,184 points were evaluated for the sensitivity analysis. (B) Second-order effects across the entire input domain (Table 1). A second-order effect captures the pairwise interaction between two parameters. Sobol indices with a 95% confidence interval that does not overlap zero are highlighted with a pink border. The largest second-order effect is emphasized with a bold pink border. (C) Predicted outbreak probabilities with varying “average infectivity” and “average mobility” parameters (i.e., the two parameters with the largest second-order effect, see panel B). Other parameters were fixed at default values (Table 1).

**Figure S4.**
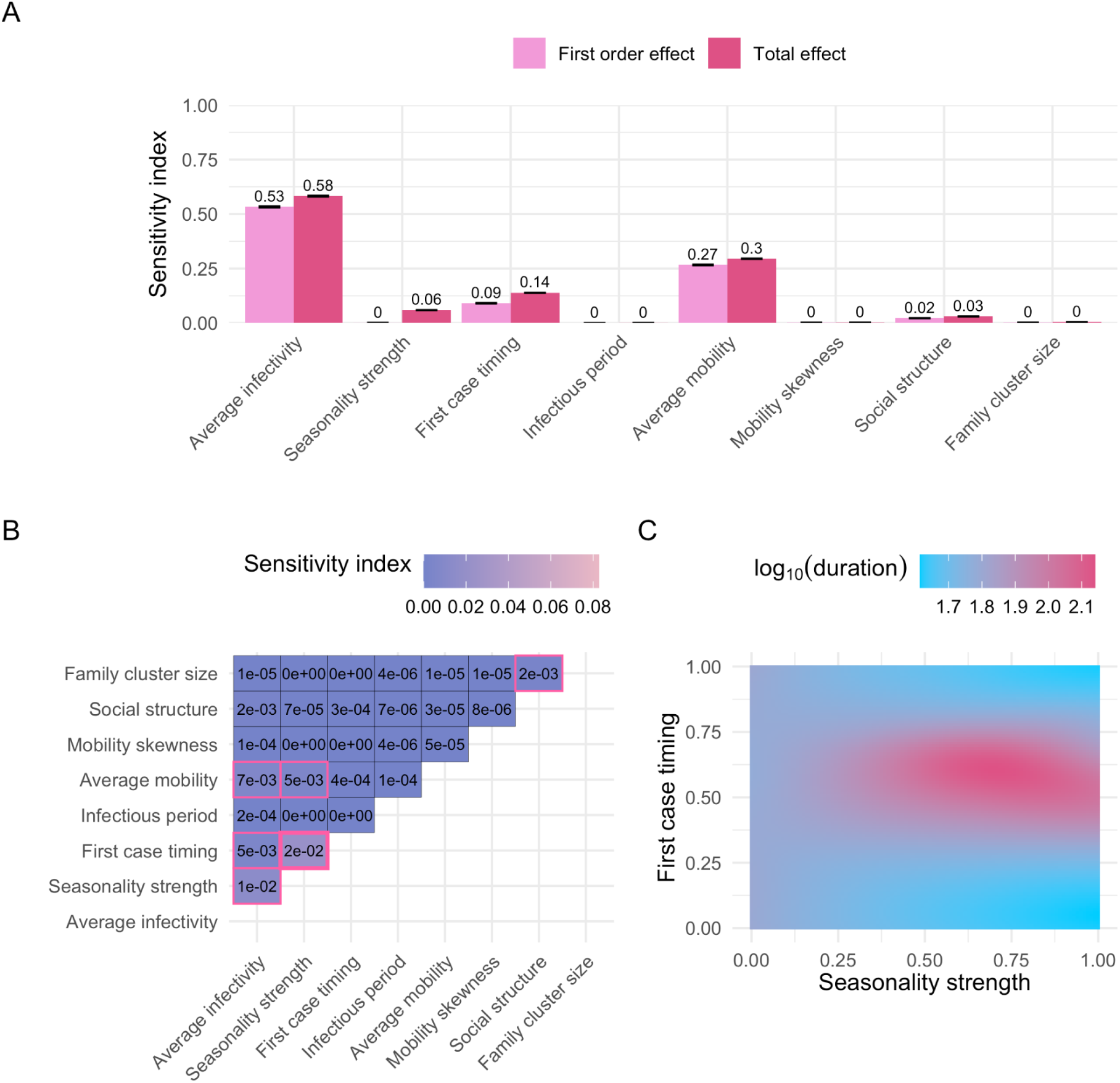
Sobol sensitivity analysis, log_10_-transformed duration. (A) First-order and total effects across the entire input domain (Table 1). The first-order effect describes the impact of a single parameter on the model output (log_10_(duration)), while the total effect accounts for all interactions involving one or more parameters. Error bars represent the 95% confidence intervals of the sensitivity index estimates. A total of 9,437,184 points were evaluated for the sensitivity analysis. (B) Second-order effects across the entire input domain (Table 1). A second-order effect captures the pairwise interaction between two parameters. Sobol indices with a 95% confidence interval that does not overlap zero are highlighted with a pink border. The largest second-order effect is emphasized with a bold pink border. (C) log_10_(duration) predictions with varying “seasonality strength” and “first case timing” parameters (i.e., the two parameters with the largest second-order effect, see panel B). Other parameters were fixed at default values (Table 1).

**Figure S5.**
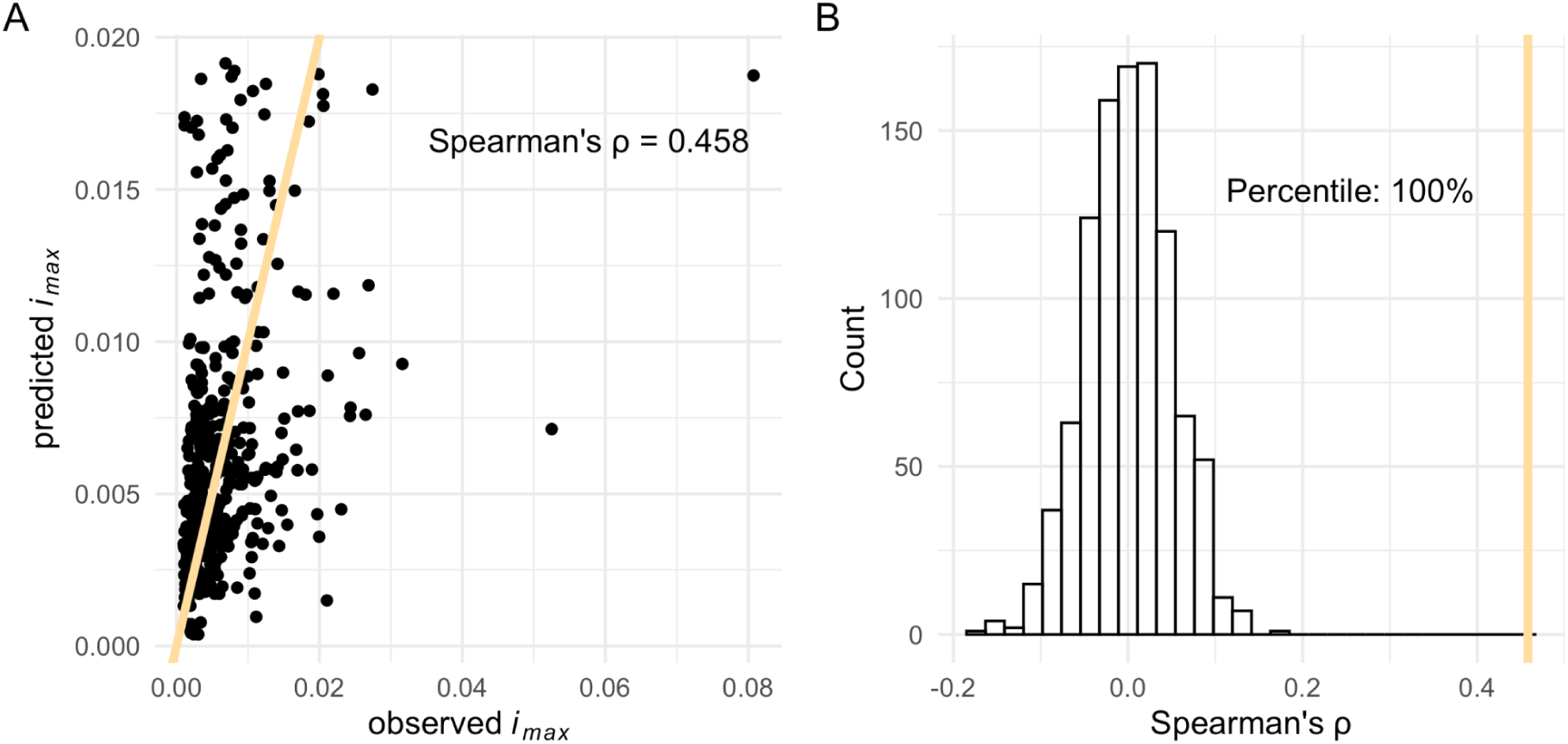
(A) Observed vs. predicted maximum incidence (*i_max_*) for empirical epidemic outbreaks (N = 449). The yellow line represents the identity line (*x = y*). (B) Distribution of Spearman correlation coefficients between observed and predicted *i_max_* from 1,000 permutations, where both the onset and municipality of the 449 epidemics were randomized. The actual observed correlation coefficient is shown as a vertical yellow line.

### Supplementary tables

**Table S1.** Summary statistics describing the 250 parameter combinations with the lowest root mean squared errors from the parameter exploration with the Gaussian Process. These combinations represent the best-fitting sets of parameters for matching observed and predicted dengue maximum incidences across municipalities. IQR = Interquartile Range.

| Parameter | Min | Mean | Max | IQR |
| --- | --- | --- | --- | --- |
| Seasonality strength | 6e-5 | 0.05 | 0.21 | 0.05 |
| First case timing | 0.00 | 0.50 | 1.00 | 0.32 |
| Infectious period | 4.00 | 5.02 | 6.00 | 0.97 |
| Average mobility | 1.00 | 2.51 | 5.00 | 1.67 |
| Mobility skewness | 0.05 | 0.48 | 0.95 | 0.40 |
| Social structure | 0.02 | 0.63 | 0.99 | 0.53 |
| Family cluster size | 1.16 | 11.17 | 19.90 | 8.71 |
| Scaling factor | 0.01 | 0.03 | 0.10 | 0.01 |

## Notes

### Competing Interest Statement

The authors have declared no competing interest.

### Funding Statement

This project has received funding from the European Unions Horizon2020 research and innovation program under the Marie Sklodowska-Curie grant agreement No. 101025586. PWM was supported by the National Institutes of Health under award R35GM152242.

### Summary of Updates

The title, introduction, and discussion were revised to clarify the purpose of the paper: exploring the potential of Gaussian Process emulation in epidemiological modeling. Main Analysis & Results have not changed.

## References

1. Grimm V, Berger U, Bastiansen F, Eliassen S, Ginot V, Giske J, et al. A standard protocol for describing individual-based and agent-based models. Ecol Modell. 2006;198: 115–126.

2. Bershteyn A, Gerardin J, Bridenbecker D, Lorton CW, Bloedow J, Baker RS, et al. Implementation and applications of EMOD, an individual-based multi-disease modeling platform. Pathog Dis. 2018;76. doi:10.1093/femspd/fty059

3. de Lima TFM, Lana RM, de Senna Carneiro TG, Codeço CT, Machado GS, Ferreira LS, et al. DengueME: A Tool for the Modeling and Simulation of Dengue Spatiotemporal Dynamics. Int J Environ Res Public Health. 2016;13. doi:10.3390/ijerph13090920

4. Hladish TJ, Pearson CAB, Toh KB, Rojas DP, Manrique-Saide P, Vazquez-Prokopec GM, et al. Designing effective control of dengue with combined interventions. Proc Natl Acad Sci U S A. 2020;117: 3319–3325.

5. Perkins TA, Reiner RC Jr, España G, ten Bosch QA, Verma A, Liebman KA, et al. An agent-based model of dengue virus transmission shows how uncertainty about breakthrough infections influences vaccination impact projections. PLoS Comput Biol. 2019;15: e1006710.

6. Smith NR, Trauer JM, Gambhir M, Richards JS, Maude RJ, Keith JM, et al. Agent-based models of malaria transmission: a systematic review. Malar J. 2018;17: 299.

7. Xu P, Liang S, Hahn A, Zhao V, Lo WT ‘jack’, Haller BC, et al. e3SIM: epidemiological-ecological-evolutionary simulation framework for genomic epidemiology. bioRxiv. 2024. doi:10.1101/2024.06.29.601123

8. Perkins TA, Garcia AJ, Paz-Soldán VA, Stoddard ST, Reiner RC Jr, Vazquez-Prokopec G, et al. Theory and data for simulating fine-scale human movement in an urban environment. J R Soc Interface. 2014;11: 20140642.

9. Sobol′ IM. Global sensitivity indices for nonlinear mathematical models and their Monte Carlo estimates. Math Comput Simul. 2001;55: 271–280.

10. Hethcote HW. The mathematics of infectious diseases. SIAM Rev Soc Ind Appl Math. 2000;42: 599–653.

11. Sacks J, Welch WJ, Mitchell TJ, Wynn HP. Design and Analysis of Computer Experiments. Stat Sci. 1989;4: 409–423.

12. Paleyes A, Mahsereci M, Lawrence ND. Emukit: A Python toolkit for decision making under uncertainty. Proc of the Python in Science Conf. 2023.

13. Matheron G. Principles of geostatistics. Econ Geol. 1963;58: 1246–1266.

14. Krige DG. A Statistical Approach to Some Basic Mine Valuation Problems on the Witwatersrand. Journal of the Southern African Institute of Mining and Metallurgy. 1951;52: 119–139.

15. Bower RG, Goldstein M, Vernon I. Galaxy Formation: a Bayesian Uncertainty Analysis. Bayesian Anal. 2010;5: 619–670.

16. Champer SE, Oakes N, Sharma R, García-Díaz P, Champer J, Messer PW. Modeling CRISPR gene drives for suppression of invasive rodents using a supervised machine learning framework. PLoS Comput Biol. 2021;17: e1009660.

17. Mubangizi M, Andrade-Pacheco R, Smith M, Quinn J, Lawrence ND. Malaria surveillance with multiple data sources using Gaussian process models. Proceedings of the 1st International Conference on the Use of Mobile ICT in Africa 2014. 2014. Available: https://inverseprobability.com/publications/files/malaria-surveillance.pdf

18. Nguyen Q. Bayesian Optimization in Action. Shelter Island: Manning Publications; 2023.

19. Gardner JR, Pleiss G, Weinberger KQ, Bindel D, Wilson AG. Gpytorch: Blackbox matrix-matrix gaussian process inference with gpu acceleration. Proceedings of the 32nd International Conference on Neural Information Processing Systems. Reed Hook: Curran Associates Inc.; 2018. pp. 7587–7597.

20. Gething PW, Noor AM, Gikandi PW, Ogara EAA, Hay SI, Nixon MS, et al. Improving imperfect data from health management information systems in Africa using space-time geostatistics. PLoS Med. 2006;3: e271.

21. Bhatt S, Weiss DJ, Cameron E, Bisanzio D, Mappin B, Dalrymple U, et al. The effect of malaria control on Plasmodium falciparum in Africa between 2000 and 2015. Nature. 2015;526: 207–211.

22. Johnson LR, Gramacy RB, Cohen J, Mordecai E, Murdock C, Rohr J, et al. Phenomenological forecasting of disease incidence using heteroskedastic Gaussian processes: A dengue case study. Ann Appl Stat. 2018;12: 27–66.

23. Albinati J, Meira W, Pappa GL. An Accurate Gaussian Process-Based Early Warning System for Dengue Fever. 2016 5th Brazilian Conference on Intelligent Systems (BRACIS). IEEE; 2016. pp. 43–48.

24. Sawe SJ, Mugo R, Wilson-Barthes M, Osetinsky B, Chrysanthopoulou SA, Yego F, et al. Gaussian process emulation to improve efficiency of computationally intensive multidisease models: a practical tutorial with adaptable R code. BMC Med Res Methodol. 2024;24: 26.

25. Andrianakis I, Vernon IR, McCreesh N, McKinley TJ, Oakley JE, Nsubuga RN, et al. Bayesian history matching of complex infectious disease models using emulation: a tutorial and a case study on HIV in Uganda. PLoS Comput Biol. 2015;11: e1003968.

26. Reiker T, Golumbeanu M, Shattock A, Burgert L, Smith TA, Filippi S, et al. Emulator-based Bayesian optimization for efficient multi-objective calibration of an individual-based model of malaria. Nat Commun. 2021;12: 7212.

27. Masserey T, Lee T, Golumbeanu M, Shattock AJ, Kelly SL, Hastings IM, et al. The influence of biological, epidemiological, and treatment factors on the establishment and spread of drug-resistant Plasmodium falciparum. eLife. 2022;11. doi:10.7554/eLife.77634

28. Golumbeanu M, Yang G-J, Camponovo F, Stuckey EM, Hamon N, Mondy M, et al. Leveraging mathematical models of disease dynamics and machine learning to improve development of novel malaria interventions. Infect Dis Poverty. 2022;11: 61.

29. Burgert L, Reiker T, Golumbeanu M, Möhrle JJ, Penny MA. Model-informed target product profiles of long-acting-injectables for use as seasonal malaria prevention. Boëte C, editor. PLOS Glob Public Health. 2022;2: e0000211.

30. Johansson MA, Apfeldorf KM, Dobson S, Devita J, Buczak AL, Baugher B, et al. An open challenge to advance probabilistic forecasting for dengue epidemics. Proc Natl Acad Sci U S A. 2019;116: 24268–24274.

31. Perrin A, Glaizot O, Christe P. Worldwide impacts of landscape anthropization on mosquito abundance and diversity: A meta-analysis. Glob Chang Biol. 2022;28: 6857–6871.

32. Paz-Bailey G, Adams LE, Deen J, Anderson KB, Katzelnick LC. Dengue. Lancet. 2024;403: 667–682.

33. Gubler DJ. Dengue and dengue hemorrhagic fever. Clin Microbiol Rev. 1998;11: 480–496.

34. Clarke J, Lim A, Gupte P, Pigott DM, van Panhuis WG, Brady OJ. A global dataset of publicly available dengue case count data. Sci Data. 2024;11: 296.

35. Siraj AS, Rodriguez-Barraquer I, Barker CM, Tejedor-Garavito N, Harding D, Lorton C, et al. Spatiotemporal incidence of Zika and associated environmental drivers for the 2015-2016 epidemic in Colombia. Sci Data. 2018;5: 180073.

36. Freitas LP, Carabali M, Yuan M, Jaramillo-Ramirez GI, Balaguera CG, Restrepo BN, et al. Spatio-temporal clusters and patterns of spread of dengue, chikungunya, and Zika in Colombia. PLoS Negl Trop Dis. 2022;16: e0010334.

37. Stoddard ST, Forshey BM, Morrison AC, Paz-Soldan VA, Vazquez-Prokopec GM, Astete H, et al. House-to-house human movement drives dengue virus transmission. Proc Natl Acad Sci U S A. 2013;110: 994–999.

38. Johansson MA, Dominici F, Glass GE. Local and global effects of climate on dengue transmission in Puerto Rico. PLoS Negl Trop Dis. 2009;3: e382.

39. Wesolowski A, Qureshi T, Boni MF, Sundsøy PR, Johansson MA, Rasheed SB, et al. Impact of human mobility on the emergence of dengue epidemics in Pakistan. Proc Natl Acad Sci U S A. 2015;112: 11887–11892.

40. Reiner RC Jr, Stoddard ST, Scott TW. Socially structured human movement shapes dengue transmission despite the diffusive effect of mosquito dispersal. Epidemics. 2014;6: 30–36.

41. Moore TC, Brown HE. Estimating Aedes aegypti (Diptera: Culicidae) Flight Distance: Meta-Data Analysis. J Med Entomol. 2022;59: 1164–1170.

42. Zahid MH, Van Wyk H, Morrison AC, Coloma J, Lee GO, Cevallos V, et al. The biting rate of Aedes aegypti and its variability: A systematic review (1970-2022). PLoS Negl Trop Dis. 2023;17: e0010831.

43. Shragai T, Pérez-Pérez J, Del Pilar Quimbayo-Forero M, Rojo R, Harrington LC, Rúa-Uribe G. Distance to public transit predicts spatial distribution of dengue virus incidence in Medellín, Colombia. Sci Rep. 2022;12: 8333.

44. Rasmussen CE, Williams CKI. Gaussian processes for machine learning. Cambridge, USA: MIT Press; 2005.

45. Gutierrez-Barbosa H, Medina-Moreno S, Zapata JC, Chua JV. Dengue infections in Colombia: Epidemiological trends of a hyperendemic country. Trop Med Infect Dis. 2020;5: 156.

46. Bhatt S, Gething PW, Brady OJ, Messina JP, Farlow AW, Moyes CL, et al. The global distribution and burden of dengue. Nature. 2013;496: 504–507.

47. Kraemer MU, Sinka ME, Duda KA, Mylne AQN, Shearer FM, Barker CM, et al. The global distribution of the arbovirus vectors Aedes aegypti and Ae. albopictus. Elife. 2015;4: e08347.

48. Nelson A. Estimated travel time to the nearest city of 50,000 or more people in year 2000. European Commission - Joint Research Centre - Forest Resources and Climate Unit. [cited 18 Mar 2024]. Available: https://forobs.jrc.ec.europa.eu/gam

49. Thai KTD, Anders KL. The role of climate variability and change in the transmission dynamics and geographic distribution of dengue. Exp Biol Med. 2011;236: 944–954.

50. Nordhaus WD. Geography and macroeconomics: new data and new findings. Proc Natl Acad Sci U S A. 2006;103: 3510–3517.

51. Khan MB, Yang Z-S, Lin C-Y, Hsu M-C, Urbina AN, Assavalapsakul W, et al. Dengue overview: An updated systemic review. J Infect Public Health. 2023;16: 1625–1642.

52. Magori K, Legros M, Puente ME, Focks DA, Scott TW, Lloyd AL, et al. Skeeter Buster: a stochastic, spatially explicit modeling tool for studying Aedes aegypti population replacement and population suppression strategies. PLoS Negl Trop Dis. 2009;3: e508.

53. González MC, Hidalgo CA, Barabási A-L. Understanding individual human mobility patterns. Nature. 2008;453: 779–782.

54. Song C, Koren T, Wang P, Barabási A-L. Modelling the scaling properties of human mobility. Nature Phys. 2010;6: 818–823.

55. Tatem AJ, Huang Z, Das A, Qi Q, Roth J, Qiu Y. Air travel and vector-borne disease movement. Parasitology. 2012;139: 1816–1830.

56. Bonilla EV, Chai KM, Williams CKI. Multi-task Gaussian Process Prediction. Proceedings of the 20th International Conference on Neural Information Processing Systems. Red Hook: Vancouver, British Columbia, Canada; 2007. pp. 153–160.

57. Balandat M, Karrer B, Jiang DR, Daulton S, Letham B, Wilson AG, et al. BOTORCH: a framework for efficient monte-carlo Bayesian optimization. Proceedings of the 34th International Conference on Neural Information Processing Systems. Red Hook, NY, USA: Curran Associates Inc.; 2020. pp. 21524–21538.

58. Schrama M, Hunting ER, Beechler BR, Guarido MM, Govender D, Nijland W, et al. Human practices promote presence and abundance of disease-transmitting mosquito species. Sci Rep. 2020;10: 13543.

59. Thongsripong P, Hyman JM, Kapan DD, Bennett SN. Human-mosquito contact: A missing link in our understanding of mosquito-borne disease transmission dynamics. Ann Entomol Soc Am. 2021;114: 397–414.

60. Wei Xiang BW, Saron WAA, Stewart JC, Hain A, Walvekar V, Missé D, et al. Dengue virus infection modifies mosquito blood-feeding behavior to increase transmission to the host. Proc Natl Acad Sci U S A. 2022;119: e2117589119.

61. Stoddard ST, Morrison AC, Vazquez-Prokopec GM, Paz Soldan V, Kochel TJ, Kitron U, et al. The role of human movement in the transmission of vector-borne pathogens. PLoS Negl Trop Dis. 2009;3: e481.

62. Grimm V, Railsback SF, Vincenot CE, Berger U, Gallagher C, DeAngelis DL, et al. The ODD protocol for describing agent-based and other simulation models: A second update to improve clarity, replication, and structural realism. J Artif Soc Soc Simul. 2020;23: 7.

63. Paszke A, Gross S, Massa F, Lerer A, Bradbury J, Chanan G, et al. PyTorch: An Imperative Style, High-Performance Deep Learning Library. Proceedings of the 33rd International Conference on Neural Information Processing Systems. Red Hook, NY, USA: Curran Associates Inc.; 2019. pp. 8024–8035.

64. Saltelli A, Ratto M, Andres T, Campolongo F, Cariboni J, Gatelli D, et al. Global Sensitivity Analysis: The Primer. John Wiley & Sons; 2008.

65. Herman J, Usher W. SALib: An open-source Python library for Sensitivity Analysis. The Journal of Open Source Software. 2017;2. doi:10.21105/joss.00097

66. Cavany SM, España G, Vazquez-Prokopec GM, Scott TW, Perkins TA. Pandemic-associated mobility restrictions could cause increases in dengue virus transmission. PLoS Negl Trop Dis. 2021;15: e0009603.

67. Mejía-Jurado E, Echeverry-Cárdenas E, Aguirre-Obando OA. Potential current and future distribution for Aedes aegypti and Aedes albopictus in Colombia: important disease vectors. Biol Invasions. 2024;26: 2119–2137.

68. United Nations Office For The Coordination of Humanitarian Affairs. Colombia - Subnational Administrative Boundaries. 20 Apr 2020 [cited 22 Feb 2024]. Available: https://data.humdata.org/dataset/cod-ab-col

69. Helwig NE. Multiple and Generalized Nonparametric Regression. London: SAGE Publications, Inc.; 2020.

70. R Core Team. R: A Language and Environment for Statistical Computing. Vienna, Austria: R Foundation for Statistical Computing; 2022. Available: https://www.R-project.org/

